# Expert Discrimination of AI-Generated versus Authentic Radiologic Images: A Multimodal, Pre-Registered Visual Turing Test

**DOI:** 10.64898/2026.06.25.26356440

**Authors:** Yoojin Nam, Taein An, Sung Il Hwang, Hong Jang, Chawoong Jeon, Ji Won Jeong, Juhyun Jeong, Dong Yeong Kim, So Yeon Kim, Sungkwan Kim, Yangwon Kim, Kwang Hwi Lee, Hee Sang Oh, Joon Hyung Park, Minkook Seo, Yongsik Sim, Jeong Min Song, Seunghyun Song, Hee Mang Yoon, the MeducAI Reader Study Group, Pa Hong, Namkug Kim

**Affiliations:** Department of Radiology, Samsung Changwon Hospital, Sungkyunkwan University School of Medicine, Changwon, Republic of Korea; Department of Convergence Medicine, University of Ulsan College of Medicine, Asan Medical Center, Seoul, Republic of Korea; Department of Radiology and Research Institute of Radiology, University of Ulsan College of Medicine, Asan Medical Center, Seoul, Republic of Korea; Department of Radiology, Hallym University Dongtan Sacred Heart Hospital, Hwaseong, Republic of Korea; Department of Radiology, Seoul National University Bundang Hospital, Seongnam, Republic of Korea; Department of Radiology, Gimcheon Jeil Hospital, Gimcheon, Republic of Korea; Department of Radiology, Kyung Hee University Hospital, Kyung Hee University College of Medicine, Seoul, Republic of Korea; Department of Radiology, Ansan Hospital, Korea University School of Medicine, Ansan, Republic of Korea; Centum Breast and Thyroid Clinic, Busan, Republic of Korea; Department of Radiology and the Research Institute of Radiological Science, Gangnam Severance Hospital, Yonsei University College of Medicine, Seoul, Republic of Korea; Department of Radiology, Samsung Medical Center, Sungkyunkwan University School of Medicine, Seoul, Republic of Korea; Department of Radiology, Massachusetts General Hospital, Boston, Massachusetts, United States of America

## Abstract

**Background:** Frontier text-to-image models can synthesise radiologic images of high realism, raising the question of whether expert radiologists can serve as a provenance safeguard for the medical image record.

**Methods:** We conducted a prospective, pre-registered visual Turing test in which 60 invited Korean board-certified radiology faculty and trainees judged authentic (teaching-repository) and AI-generated radiologic images from a locked pool of 241 displayable cells (82 entities; nine subspecialties; six modalities; 60 readers x 60 trials = 3,600 reader–image observations) produced by two contemporary commercial generators. The primary endpoint was the confidence-weighted, reader-averaged multi-reader multi-case area under the curve for AI versus authentic images, conditional on the locked image pool; the key secondary endpoint was the Faculty-minus-Junior difference under a two one-sided tests equivalence framework. The pre-specified statistical analysis plan was registered on the Open Science Framework before data lock.

**Findings:** All 60 readers completed the test. The pooled confidence-weighted area under the curve was 0.71 (95% CI, 0.69 to 0.74), above the null value of 0.5 but within the pre-specified modest tier (0.60 to 0.75). The Faculty-minus-Junior contrast was 0.04 (95% CI, −0.02 to 0.10), including zero, and the two one-sided tests established equivalence within the +/−0.10 margin. No reader stratum and no pre-specified sensitivity analysis reached the deployable-classifier threshold (area under the curve >= 0.75).

**Interpretation:** In this single-country cohort, expert radiologists distinguished frontier-generated from authentic radiologic images only modestly, without a meaningful expertise gradient (equivalence within +/−0.10) and with no reader stratum reaching a standalone provenance safeguard. These findings support radiology AI-literacy training and pipeline-level provenance safeguards rather than reliance on reader judgment, and warrant retesting in an independent reader cohort.

**Funding:** This research was supported by a grant of the Korea Health Technology R&D Project through the Korea Health Industry Development Institute (KHIDI), funded by the Ministry of Health & Welfare, Republic of Korea (grant number: RS-2025-02213531).

## Introduction

Modern text-to-image generators now produce radiologic images of sufficient realism that they can be difficult to distinguish from authentic clinical examinations^1–3^. As these images become more convincing, they are reaching the channels through which the medical image record is assembled and trusted: radiology teaching files, figures in the peer-reviewed literature, and image-based clinical communication. The credibility of that record rests on the assumption that a radiologic image depicts a real patient examination, and the inappropriate handling of images is already a recognised threat to the integrity of the published biomedical record^4^. This is no longer hypothetical for artificial intelligence: in 2026 a New England Journal of Medicine clinical-image case was retracted after the authors used an AI tool to alter the submitted image, an alteration identified only by post-publication reader scrutiny rather than by any systematic provenance check^5^. Generative models sharpen the threat further, because they do not merely alter an existing image but synthesise a plausible one whole from a text prompt^6,7^. The decision-relevant question is therefore not only whether such images can be generated, but whether the human experts who curate teaching material, review manuscripts, and interpret images can themselves serve as a safeguard against synthetic provenance. This question arose directly in our own practice: while developing AI-assisted examination-preparation material, faculty radiologists judged that pipeline-generated images carried subtle departures from genuine examinations and could distort the perceptual learning on which radiological expertise depends, which led our group to move from generated radiologic images toward infographic-based teaching content. Prior work from our group characterised the factual content accuracy of such pipelines; the visual realism of their images, and radiologists’ ability to recognise it, was not formally evaluated and is the focus of the present study.

A recent study in *Radiology* evaluated GPT-4o-generated and authentic chest and bone radiographs with seventeen radiologists and found that only 41% of blinded readers spontaneously identified the synthetic images^1^; that work is rigorous but bounded by design to a single generator, a single modality, raw accuracy without confidence weighting, and a narrow subspecialty range. A 2026 study in *npj Digital Medicine* extended multimodal large-language-model evaluation to in-training examination items but excluded image-based questions because current models cannot yet reliably generate authentic, clinically integrated images^8^, which is the very capability a provenance-discrimination study must probe; and a separate benchmark used eye-tracking to characterise where clinicians look when judging AI-generated chest radiographs but framed the problem as gaze realism rather than as an expertise-stratified detection estimate^9^. That readers may over-trust AI-assisted outputs is well documented in radiology^10^, which is precisely why the decision-relevant quantity is the human reader’s provenance-discrimination ability rather than the generator’s pixel-level fidelity alone. Three gaps therefore remain in the current evidence: no study has evaluated two contemporary commercial generators within a single locked frame against the same authentic entities; none has reported a confidence-weighted multi-reader multi-case area under the curve with a pre-registered expertise contrast under an equivalence framework, the structure required to translate detection performance into deployment-relevant thresholds and to test, rather than assume, an expertise gradient; and none has reported a prespecified analysis of which provenance cues are associated with correct detection, which is what is needed to judge whether reliable safeguards could be built on human reading at all.

We conducted a prospective, pre-registered visual Turing test of radiologic image provenance discrimination on a locked pool of 241 displayable cells spanning 82 entities across nine subspecialties and six modalities, with 60 invited Korean board-certified radiology faculty and trainees stratified into three expertise tiers (60 readers x 60 trials = 3,600 reader–image observations). The study tests three pre-registered hypotheses. The primary hypothesis (H1) is that radiology readers discriminate AI-generated from authentic radiologic images above chance, evaluated as the confidence-weighted multi-reader multi-case area under the receiver operating characteristic curve for the pooled AI arm (Nano Banana 2 and gpt-image-2) against the authentic arm, tested against a null value of 0.5^11,12^, and interpreted against three pre-specified substantive tiers (near-chance, modest-above-chance, and deployment-relevant) rather than against the null alone. The key secondary hypothesis (H2) is that discrimination differs by expertise level, tested as a pre-specified Faculty-minus-Junior difference in the area under the curve under a two-one-sided-tests equivalence framework with a margin of plus or minus 0.10^13–15^, so that a null contrast is itself an interpretable finding rather than an absence of evidence. The prespecified secondary hypothesis (H3-Q3) is that the provenance cue a reader reports is associated with trial-level accuracy within each expertise stratum, the analysis needed to identify which reader-reported cues are associated with correct detection. H3-Q3 was prespecified before broadcast under a no-data-seen attestation lodged at the Open Science Framework^16^.

The study has two complementary aims. As a question of image-record integrity, it asks whether expert radiologists can reliably distinguish synthetic from authentic radiologic images, and therefore whether human reading can serve as a provenance safeguard for teaching files, journal figures, and clinical image communication as generative models mature. As a question of training, it asks whether such discrimination depends on expertise and which provenance cues are associated with accurate detection and could inform radiology AI-literacy curricula; a demonstration of curriculum effectiveness is delegated to a parallel companion study in radiology medical education from our group^17^. Secondary implications for editorial figure-provenance auditing and journal anti-fabrication screening are exploratory hypotheses rather than uses this design validates: should expert discrimination reach a pre-specified threshold (an area under the curve of at least 0.75 with a lower 95% confidence bound of at least 0.70, jointly satisfied by the primary pool and a quality-restricted sensitivity analysis), this would identify provenance screening as a direction warranting dedicated classifier development and external clinical-spectrum validation, not as a safeguard the present human-reader study licenses. The study does not claim to validate any deployed image-provenance classifier; consistent with this scope, a null or near-chance result among experts is itself an informative finding, indicating that human reading is a limited safeguard and that systematic, technology-level provenance protections and AI-literacy training are needed.

## Methods

### 1. Study design and overview

This was a prospective, pre-registered, multi-institutional visual Turing test of radiologic image provenance discrimination (readers recruited from more than 30 institutions nationwide under single-site institutional-review-board oversight). Board-certified radiology faculty and radiology trainees were asked to judge, for each of a series of normalised radiologic images, whether the image was authentic or generated by a contemporary text-to-image model, and to report their confidence and the perceptual cue underlying their judgement. The synthetic arm was produced by the upstream image-generation pipeline previously described by our group for radiology teaching-material synthesis^18^, using two vendor-distributed generators frozen at a fixed model snapshot; the pipeline implementation is publicly archived (https://github.com/Yoojin-nam/MeducAI, Apache-2.0, deposited at Zenodo with a versioned DOI at publication) and the frozen prompts, model identifiers, and per-image provenance are reported self-containedly in Supplementary Methods S3, so the synthetic-arm generation is reproducible independently of that companion paper’s publication status. Each reader evaluated 60 trials drawn from a locked image pool of 241 displayable cells (83 authentic, 78 from the Nano Banana 2 arm, and 80 from the gpt-image-2 arm) spanning 82 distinct radiologic entities, presented as 83 entity-modality units (one breast entity appears in both mammography and ultrasound). Readers were blinded to image provenance and to the proportion of authentic versus synthetic images in their assignment.

The study was preregistered at the Open Science Framework before any reader was exposed to the task (registration of record https://osf.io/zrwy8;^16^) and the full statistical analysis plan was locked, with the final reader-image assignment hash, before broadcast. The protocol and the analysis plan were approved by the Institutional Review Board of Samsung Changwon Hospital, Sungkyunkwan University School of Medicine (File No. SCMC IRB 2025-12-020; the visual Turing test was incorporated under amendment 2025-12-020-002, approved on 8 May 2026 by expedited review, with an approval period of 7 January 2026 to 6 January 2027; principal investigator Pa Hong); reader participation was voluntary under electronic informed consent, and reader responses were collected through token-authenticated web forms. The reader task was broadcast on 2026-06-05 at 09:00 Korea Standard Time, with a planned 14-day response window (closing 2026-06-19 23:59 KST) and a pre-registered reminder-based grace window of up to a further seven days (to 2026-06-26 23:59 KST) for any reader who had not completed. Because all 60 readers completed the task before the 2026-06-19 deadline, the grace window was not invoked, and the data were locked on 2026-06-20, after the registered response window had closed. The pre-registered extension rule keys on completion status only, never on response content (see the Deviations and amendment policy subsection).

### 2. Synthetic image generation protocol

#### Models and freeze policy

Two image-generation models were used: Nano Banana 2 (Gemini 3.1 Flash Image, Google DeepMind, 2026) and gpt-image-2 (OpenAI, 2026). Both model snapshots were frozen at frame lock (2026-04-28, 23:59 KST), and a per-record model-snapshot identifier was preserved for every generated image. The two models were chosen to span the frontier and the next-generation tier of vendor-distributed image generation at the time of frame lock, enabling a within-frame comparison of synthetic-image reliability across providers without altering the locked entity matrix.

#### Prompt design framework

A structured five-block prompt architecture common to both models was used: a modality-fidelity block (modality, view or sequence, technical parameters); an anatomical-region block (organ system, organ, subregion, laterality, landmarks, forbidden structures, adjacency rules); a finding block (key-finding keywords, conspicuity calibration, severity range); a technical-quality block (PACS-realistic grayscale, soft edges, no overlays or text); and a negative prompt with a post-generation compliance check. Each per-image specification carried a constraint block providing entity-specific anatomical constraints that the prompt was required to honour, including organ topology, forbidden adjacent structures, and view-exclusion hints.

#### Iterative prompt refinement

The prompt-improvement procedure comprised three distinct phases. In Phase A (2025-12-28 to 2026-01-04), prompt improvement for the text-generation stages S1 (entity-table generation) and S2 (board-style card generation) was implemented as a closed-loop pipeline in which the pipeline’s own LLM-as-a-judge validation stage (S5) audited each generated entity table and card and emitted a structured per-issue record carrying an issue code, a severity flag, a recommended fix target, a draft natural-language patch hint, and a blocking-error flag; these records were aggregated by a deterministic transform into a prioritised backlog, the lead investigator committed prompt edits for each blocking and selected high-frequency issue, and the resulting prompts were re-validated on a held-out development cohort. Three rounds were executed (S5R0 baseline, S5R1, S5R2); the S5R2 prompts are the operative text prompts for this study. In Phase B (2026-04-08 to 2026-04-22), prompt improvement for the Stage S4_EXAM image-generation prompt was conducted by lead-investigator human review of generated images on a nine-entity development set held independent of the locked study frame, so that prompt selection could not be tuned to study outcomes; the image prompt advanced through four manually authored versions (v13 to v16, the last frozen for full-frame generation). In Phase C (2026-04-22), a single model-specific variant for gpt-image-2, denoted v16.1, re-used v16 verbatim as a scaffold and introduced four targeted edits informed by an independent failure-mode analysis of an earlier-generation image generator reported by Tordjman and colleagues^1^, targeting pathology mislocalisation, disease erasure, view confusion, and view-exclusion violation; this was a one-shot transfer rather than a refinement loop. Cumulative per-version prompt diffs are provided in the Supplement.

#### Frozen prompts and provenance

The final locked prompts were, for the Nano Banana 2 arm, the v16 system prompt (SHA-256-16 `777ef1444a88ae2d`) and user template (`1212abd0b8cc5f90`), and for the gpt-image-2 arm, the v16.1 system prompt (`72683f8aab802495`) and user template (`4c2cce116622b7bc`). Every per-image manifest record carries the system and user prompt hashes, and a manifest-level frame hash allows the run-time entity matrix to be reproduced bit-for-bit.

#### Image generation execution

For Nano Banana 2 (v16), images were generated through the Google Generative AI SDK at temperature 0.2 with a single pass per entity. For gpt-image-2 (v16.1), images were generated through the OpenAI Batch API with seed 42, quality “high”, and a single combined prompt per entity. Per-image telemetry (call identifier, request timestamps, finish reason, prompt feedback, safety ratings, retry attempts, and image SHA-256) was stored for every record. The full-frame Nano Banana 2 run completed on 2026-04-29 (manifest SHA-256 `1a3bd058…751984`), with all entities saved and no moderation refusals; the full-frame gpt-image-2 run yielded the remaining entities together with a small number of vendor-side moderation refusals concentrated in anatomically sensitive (breast and neonatal) regions. The locked prompt was not modified to retry refused entities; refused entities are reported as policy artefacts with full safety-violation telemetry.

#### Amendment policy

Two frame amendments occurred after the prompt freeze (v23r4 and v23r5, 2026-05-01 to 2026-05-02); both substituted entities within unchanged subspecialty-by-modality cells and altered no prompt file. For each amendment the substituted entities were re-passed through the downstream stages using the byte-identical locked prompt files, and image-level SHA-256 was recomputed for each new image.

### 3. AI-mediated recursive prompt refinement: claim and scope

The improvement pipeline contained one phase meeting a defensible operational definition of recursive AI-mediated refinement (Phase A, the S5R closed loop applied to the text prompts) and two phases that did not (Phase B, image-prompt human iteration; Phase C, one-shot external-evidence transfer). The boundary of the AI-mediated component is reported explicitly. The S5R loop met four conditions of an AI-mediated recursive refinement pipeline: the issue signal was generated by a language model; the prompt-patch proposal was generated by a language model; the proposals were aggregated and prioritised by a deterministic transform; and the loop was iterated and the iterations were quantitatively compared. The loop is therefore best characterised as a semi-automated, AI-mediated, evaluator-driven prompt-refinement pipeline rather than a fully autonomous one, because a human author committed each prompt edit and the validator’s patch hint served as a structured suggestion rather than an executed change.

The loop is reported transparently, and we do not claim that it improved the pre-registered primary endpoint of the refinement experiment. In a pre-registered development experiment comparing the S5R0 and S5R2 prompts on 11 paired group identifiers, the pre-registered primary endpoint (the per-group rate of any S2 issue) decreased from a median of 24.5% to 19.0% (median paired difference −2.75 percentage points; bootstrap 95% CI −7.79 to +5.01; p = 0.66; 6 of 11 groups improved). The pre-registered expansion criterion (directional improvement in 9 of 11 groups) was not met; the S5R loop was therefore not extended to the holdout cohort, and the S5R2 prompts were frozen for production. Three image-related secondary endpoints were directionally favourable (for example, the clean-image rate improved in 8 of 11 groups, p = 0.21), although the corresponding confidence intervals also included zero. To our knowledge this is one of the first reported effect estimates for an LLM-as-a-judge-driven prompt-refinement loop on a medical-content generation pipeline, and we report the negative-but-informative result as such.

For the image prompt (Phase B), no automated critique loop was implemented; iteration v13 to v16 was performed by lead-investigator human review of development-set images, and we make no claim of AI-mediated refinement for this phase. The single edit v16 to v16.1 (Phase C) transferred an external failure-mode taxonomy into the gpt-image-2 prompt and is an evidence-based external transfer rather than a refinement loop. No fully autonomous closed-loop optimisation procedure was used at any phase, no language-model critique of development-set images was fed back into a subsequent image-prompt version, and no language-model-generated edit was committed without human review. The model identities, access modes, stochasticity controls, prompt provenance, and development-test independence of the generative components are reported against the MI-CLEAR-LLM checklist in Supplementary Methods S3, reproduced from the canonical MI-CLEAR-LLM mapping. In brief, the Nano Banana 2 image arm used temperature 0.2 and the gpt-image-2 arm used seed 42, each with a single query per entity and a per-image SHA-256 as the reproducibility token, and the frozen prompt files are identified by SHA-256 hash; the upstream S5 LLM-as-a-judge content validator that audited the entity-table (S1) and card (S2) stages used Gemini 3 Pro Preview (entity-table validation) and Gemini 3 Flash Preview (card validation) at temperature 0.2 with reasoning and retrieval enabled via the chat application-programming interface, consistent with the shared pipeline reported in the companion Paper 1 supplementary materials; its access mode, run configuration, and the development-set (held independent of the study frame) on which the refinement loop was evaluated are reported in Supplementary Methods S3.

### 4. Image-format harmonisation

All images presented to readers underwent a single operative preprocessing pipeline applied symmetrically across the authentic, Nano Banana 2, and gpt-image-2 arms: a crop-free, modality-aware normalisation that preserved diagnostic content without centre-cropping or the addition of artificial borders. Within each modality the same normalisation policy was applied to all three arms. Metadata, file path, filename, compression, and resolution cues were harmonised, and visible burned-in labels or source-identifying marks were screened during final visual quality control. This is not a post-hoc augmentation: anatomy is not altered, and images are not made artificially harder to evaluate. Width, height, and file-size distributions were inspected as a pre-dispatch descriptive quality-control step; no Kolmogorov–Smirnov-style pass/fail significance test was used as a normalisation criterion, because non-significance does not establish indistinguishability.

The public reader-facing payload exposed only opaque, token-hash-keyed assignment records — a randomised image identifier, an image URL under the normalised-asset directory, and the normalised-image hash — while the ground-truth label and image type existed exclusively in a private answer key and were joined to reader submissions on the image identifier at analysis time. This generation-provenance label is the reference standard of the study; because provenance is fixed deterministically at the moment of image creation, the reference standard is unambiguous and requires no separate adjudication, positivity cutoff, or assessor blinding. A confirmatory re-score of a small set of normalised images by the second reader (to verify that normalisation did not shift apparent realism) was deferred to the manuscript stage; should that check reveal a normalisation-induced realism shift, a preregistration deviation will be logged and a corresponding sensitivity analysis added, and otherwise it is reported as descriptive quality control only.

### 5. Image pool and frame lock

The frozen image pool was the locked image-pool manifest of 241 displayable cells across 82 distinct entities (83 authentic, 78 Nano Banana 2, and 80 gpt-image-2), with manifest SHA-256 `100ef0c0…6558749e`. The distinct-entity count is 82 — equivalently 83 entity-modality units, because one breast entity appears in both mammography and ultrasound, and 83 authentic cells; the full pool-composition lineage is enumerated in Supplementary Table S1. The pool reflects a documented assembly lineage to the locked manifest; the authentic arm received no further changes after a 19-image view/specification alignment refinement applied during pool assembly (see the Image swap audit transparency subsection). Five Nano Banana 2 cells were removed under a single prospectively locked consensus-fail rule, and two rater-confirmed clerical corrections were applied before any reader exposure (see the Quality assurance subsection); the pool-composition lineage from the original 243-cell frame to the 241-cell lock is enumerated in Supplementary Table S1. The authentic arm is sourced from the Radiopaedia teaching repository; consequently, per-image scanner model, acquisition-protocol parameters, and patient demographics are not consistently available, and the pool represents teaching-archive authentic images rather than a consecutive clinical imaging spectrum. The detectability estimand is accordingly scoped to teaching-quality authentic images, consistent with the AI-literacy intended use, and no external evaluation on an independent clinical-spectrum authentic arm is performed.

The reader-image assignment was a balanced, incomplete reader-image assignment of 60 invited readers, each assigned 60 trials, for 3,600 assignment rows, constructed so that no reader saw any entity more than once (reader-by-entity duplicate count of zero) and so that each reader’s trials comprised exactly 20 authentic, 20 Nano Banana 2, and 20 gpt-image-2 images (arm-balanced); formal block-design balance parameters were not imposed beyond the per-arm balance and no-repeat constraints. Design feasibility was verified on five of five replicate constructions, including a worst-case scenario in which all nine quality-assurance faculty also served as readers. Entities seen by a faculty rater during quality assurance were excluded from that rater’s reader assignment at the entity level, with the exclusion key being the entity itself, so that for a dual-modality entity all of its modalities are removed from that rater’s assignment.

### 6. Quality assurance and inter-rater reliability

#### Quality-assurance instrument and calibration

Every displayable synthetic cell was assessed on a six-axis instrument comprising one binary axis (medical validity) and five ordinal Likert axes (modality accuracy, main finding, additional finding, anatomical accuracy, and image quality), supplemented by a five-category texture-and-artefact checklist with per-category confidence. Before the main quality-assurance round, calibration data were collected on a 10-image, frame-external anchor set balanced across nine subspecialties and stratified by expected difficulty, with anchor stratum blinded to raters. Krippendorff’s α was computed for each axis with a 2,000-iteration bootstrap 95% CI at seed 42^19,20^. A pre-specified α launch gate (≥ 0.50) was converted, before any main quality-assurance data collection, to a reporting-only specification: the manuscript reports the α point estimate and 95% CI against the reporting tiers (operational ≥ 0.50, tentative ≥ 0.667, definitive ≥ 0.80), and substantive interpretation uses the tier system rather than the gate. Because the calibration α for several graded axes was low and imprecise at the calibration sample size, the three-tier collapsed scale (1–2 / 3 / 4–5) is the primary reporting scale for axes whose five-point α fell below 0.50. The primary endpoint of the reader study does not depend on Likert-axis agreement.

#### Second-reader inter-rater reliability

A pre-specified second faculty rater (co-principal investigator) independently scored a stratified random subset of 60 synthetic cells (seed 42; 30 Nano Banana 2 and 30 gpt-image-2; drawn from the 168 displayable cells at the v23r7r1 frame state under the constraint of a positive realism pass, invariant under the later forks), blinded to first-rater responses, using the same six-axis instrument. Inter-rater reliability is reported as the intraclass correlation coefficient (two-way mixed, single measures, absolute agreement) for the five graded axes, Cohen’s κ for the binary axis, and Fleiss’ κ for each artefact category with at least 10% prevalence, each with a 2,000-iteration bootstrap 95% CI at seed 42. An intraclass correlation of at least 0.60 on a majority of graded axes is a reporting threshold (not a launch gate) for describing agreement as acceptable^20^; the second reader is excluded from the reader cohort, and no launch decision, endpoint, or sensitivity branch is conditional on any reliability value.

#### Pre-specified pool drop rule and clerical corrections

Two source-data corrections, each initiated by the original rater confirming a clerical error in their own response, were applied before dispatch (one moving a cell’s medical-validity score from 0 to 1, retaining the cell; one moving a cell from 1 to 0); original rows were never modified, and corrections were applied as audit-traceable axis-level patches. After these corrections, a single prospectively locked consensus-fail rule removed cells satisfying both a corrected first-rater medical-validity score of 0 and a principal-investigator visual-realism assessment of fail; five Nano Banana 2 cells met this rule and were dropped from the primary pool. The rule is mechanical and admits no per-cell discretion; whether a cell falls in the drop set or the retain set is fully determined by the post-correction source-data state. To address the post-hoc-rule-writing concern transparently, the chronology of the rule relative to the upstream rater corrections and realism assessments is recorded in the lock package, and a supplementary alternative-rule table reports the drop-set size under several plausible consensus-fail predicates to demonstrate that the rule was not tuned to the observed five-cell drop set. A pre-specified statistical-analysis-plan amendment had previously included a 12-image cross-subspecialty overlap construct; a dispatch-pipeline gap omitted the overlap rows from the deployed instrument, and to preserve the data of raters who had already responded and to protect the collection deadline, the overlap construct was removed in its entirety before dispatch. Per-rater severity is therefore captured solely by the rater random intercept in the primary mixed model, and per-subspecialty means are reported with rater-cluster bootstrap confidence intervals. A response-duration (fast-click) outlier audit is reported as supplementary descriptive quality control only and does not adjust any endpoint.

### 7. Visual Turing test instrument and readers

Each trial presented one normalised image from the locked pool together with four response items. The first item was a binary provenance call (real or AI). The second item was a 1-to-5 confidence rating for that call; the binary call and its confidence together form a directional confidence-weighted score, from confident-real to confident-AI, that is the input to the confidence-weighted area-under-the-curve analysis. No indeterminate or abstain option was offered, so there are no indeterminate index-test results to handle, and the influence of low-confidence responses is examined in a pre-specified abstention-proxy sensitivity analysis. (Image modality, where it enters the correctness model as a covariate, is the known modality of the displayed image rather than a reader-reported judgment.) The third item was a single-select provenance cue from seven prespecified categories (anatomy or plausibility, modality texture or noise, pathology realism, image quality or artefact, labelling or text or metadata, gestalt, and guessed or uncertain), which is the input to the prespecified cue-by-accuracy analysis. The fourth item was an optional free-text description of the reader’s reasoning, analysed as exploratory thematic content. The reader-facing briefing did not disclose the generator identities, the exact pool composition, the per-reader arm split, or any detection-cue list; the seven cue categories were presented as neutral response options. Accordingly, the cue analysis is reported as a reader-reported selection from prespecified categories rather than as fully spontaneous, unprompted cue discovery.

Readers were recruited as a volunteer convenience series through study-enrolment and collaboration channels rather than as a consecutive or random sample, and were stratified into three expertise tiers per the locked classification: Junior (first- and second-year residents), Senior (third- and fourth-year residents), and Faculty (board-certified radiologists). Among the 60 dispatched readers, the as-recruited composition was 20 Faculty, 22 Junior, and 18 Senior, with the binding key contrast being Faculty (n = 20) versus Junior (n = 22). Each reader completed 60 trials in approximately 45 to 55 minutes; single-sitting completion was preferred but readers could resume through their authenticated link, with all responses autosaved, and within-reader duplication was prevented by upsert on a composite key of reader, trial index, and image identifier. Three named individuals who did not participate in the reader task are acknowledged but are not authors of this study and were excluded from dispatch; the co-investigator who developed the generation pipeline was likewise excluded from the reader pool to preserve evaluator–investigator independence and the no-data-seen attestation, and is retained as a named author (see the Deviations and amendment policy subsection).

### 8. Hypotheses and endpoints

Three hypotheses were pre-registered. The primary hypothesis (H1) is that radiology readers discriminate AI-generated from authentic radiologic images above chance; its endpoint is the confidence-weighted multi-reader multi-case area under the receiver operating characteristic curve for the pooled AI arm (Nano Banana 2 plus gpt-image-2) versus the authentic arm, tested against a null value of 0.5. The key secondary hypothesis (H2) is that detection performance differs by expertise level; its endpoint is the expertise-stratified, confidence-weighted area under the curve, with the pre-specified Faculty-minus-Junior difference reported with a 95% confidence interval, while the monotonic Junior-to-Senior-to-Faculty trend is treated as exploratory only. H2 is tested, not assumed, and a null Faculty–Junior contrast is a publishable finding under the framing rule described in the Manuscript framing subsection. The prespecified secondary hypothesis (H3-Q3) is that the reader-selected provenance-cue category is associated with trial-level accuracy within each expertise stratum; its endpoint is a cue-by-accuracy contingency analysis within each of the three strata, using a χ² test with continuity correction,Cramér’s V with bootstrap 95% CI, per-cue accuracy proportions with Wilson 95% CIs, and a Benjamini–Hochberg false-discovery-rate supplement. The exploratory hypothesis (H3) is that the cue-category distribution differs by expertise stratum without an accuracy join, reported as a descriptive contingency table with a χ² test and an optional taxonomy applied to the free-text responses. The cue-by-accuracy analysis was prespecified on 2026-05-23, 13 days before broadcast, with a no-data-seen attestation lodged at the preregistration.

### 9. Sample size and minimum detectable effects

Minimum detectable effects were computed under multi-reader multi-case variance assumptions (reader variance component 0.001 and reader-by-case variance component 0.0005) drawn from the diagnostic-accuracy literature^11,12,15^. The binding power statement is the as-recruited composition: 60 readers comprising 20 Faculty, 22 Junior, and 18 Senior. For the primary pooled-AUC test against 0.5, this composition retained power above 0.99 at any true area under the curve of at least 0.55. For the operative Faculty-minus-Junior contrast (Faculty n = 20 versus Junior n = 22), the minimum detectable difference at 80% power and two-sided α = 0.05 was approximately 0.038, with a difference of 0.05 detectable at about 0.92 power and a difference of 0.03 underpowered; the precision actually achieved for the equivalence test on the realized cohort is characterised by the standard error and minimum detectable difference reported below rather than by a single power figure. The cue-by-accuracy and cue-distribution analyses, operating over the full set of trial-level observations, retained power above 0.99 against an independence null at a small effect size. The originally registered design target was 65 readers (25 Junior, 25 Senior, 15 Faculty), under which the Faculty-minus-Junior minimum detectable difference was approximately 0.043; that figure is retained as the historical a priori specification, but the binding power basis for the analysis is the as-recruited 20-versus-22 contrast above.

The variance assumptions are taken from radiologic-diagnosis rather than provenance-discrimination tasks, so the Faculty–Junior minimum detectable difference is additionally reported across a pre-specified sensitivity grid of reader and reader-by-case variance components. Under this pre-specified rule, after approximately the first ten readers completed the task the empirical reader-variance component was estimated from the partial data (variance components only, with no examination of any endpoint estimate). The empirical reader-variance component lay within the pre-specified grid but implied a Faculty–Junior minimum detectable difference more than 50% larger than the design value; under the pre-specified rule this triggered a preregistration amendment, lodged before any primary-endpoint unblinding, recording the empirical estimate. On the complete cohort the standard error of the Faculty–Junior contrast was 0.031, implying a minimum detectable difference of approximately 0.09 at 80% power, versus the design value of 0.038. The Faculty–Junior difference is interpreted under a two-one-sided-tests equivalence framework^21,22^ with an equivalence margin of ±0.10, evaluated by the joint 90% confidence interval; the four reporting categories (significance and equivalence each rejected or not) are pre-specified. Because the ±0.10 margin is wider than the design minimum detectable difference, an equivalence conclusion at ±0.10 is not interpreted as excluding a smaller (0.05–0.10) expertise gradient, which the significance test addresses separately. The ±0.10 margin is justified on substantive grounds rather than as a multiple of the design minimum detectable difference: it corresponds to the upper range of routine inter-reader area-under-the-curve variability in radiology reader-performance studies, so that a difference within ±0.10 is no larger than the reader-to-reader variation already accepted in the field^15^. The empirically widened minimum detectable difference lowers the power of the equivalence test, so the equivalence result is interpreted together with this reduced power: an equivalence conclusion at ±0.10 does not exclude a smaller (0.05–0.10) expertise gradient, which the significance test addresses separately. For the observed contrast the joint 90% confidence interval (−0.01 to 0.09) lay within the ±0.10 margin, so equivalence was met; the 95% confidence interval (−0.02 to 0.10) included zero, so a significant Faculty–Junior gradient was not established. The recruitment target for Faculty was a minimum of 15 and a recommended 20, which the dispatched set met; were the completed Faculty count to fall below 12, faculty-specific estimates would be reported descriptively with confidence intervals and the H2 contrast would not be interpreted as primary inference. The final power figures are replaced at analysis time with simulation output generated under the locked frame and locked assignment matrix before data collection.

### 10. Primary analysis (H1)

For the primary endpoint, the Nano Banana 2 and gpt-image-2 arms were pooled as a single AI class. The estimand is the confidence-weighted multi-reader multi-case area under the curve for the binary task of identifying AI-generated images, pooled across all readers in the full analysis set and all images in the locked pool, with a 95% confidence interval against a null value of 0.5 at two-sided α = 0.05. Because the synthetic and authentic arms are not matched on native aspect ratio (see the Aspect-ratio cue subsection), this estimand reflects readers’ provenance discrimination together with any residual format cue; the aspect-ratio analysis bounds the magnitude of that residual contribution, and the within-aspect-ratio-stratum reading, where the arms overlap, is the de-confounded estimate. Inference uses the Obuchowski-Rockette/DeLong approach with a reader-cluster bootstrap of 2,000 iterations, computed primarily with established multi-reader multi-case software and with a DeLong confidence interval reported as a supplementary cross-check^12–14^. Model-specific areas under the curve for the Nano Banana 2 and gpt-image-2 arms are reported separately and descriptively with confidence intervals, and the contrast between the two synthetic arms is a secondary analysis reported with a 95% confidence interval and a nominal two-sided p-value that does not define the primary endpoint.

Statistical significance against 0.5 is interpreted against three pre-specified substantive tiers: a near-chance tier (0.50 to 0.60), in which discrimination is statistically detectable but not substantively meaningful and AI-image realism is essentially indistinguishable to the relevant reader stratum; a modest-above-chance tier (0.60 to 0.75), in which discrimination is real but insufficient as a standalone provenance filter, supporting the development and prospective evaluation of AI-literacy training and process safeguards; and a clinically meaningful tier (at least 0.75), in which discrimination is sufficient to support human-in-the-loop deployment subject to use-case calibration. No deployable-classifier or provenance-screening claim is made from the human-reader area under the curve at any magnitude; should discrimination reach an area under the curve of at least 0.75 with a lower 95% confidence bound of at least 0.70, jointly with the concordance criterion of the quality-restricted sensitivity (see the Sensitivity analyses subsection), this is interpreted as identifying provenance screening as a hypothesis warranting dedicated classifier development and external clinical-spectrum validation, not as a finding that licenses screening. If the model-specific areas under the curve for the two synthetic arms differ by at least 0.10 on the point estimate, the primary interpretation pivots to model-stratified reporting, with the pooled estimate retained as a supplementary summary; confidence-interval overlap is not a precondition for invoking this rule.

### 11. Key secondary analysis (H2)

The key secondary analysis reports the expertise-stratified, confidence-weighted area under the curve for the Junior, Senior, and Faculty strata, each with a 95% confidence interval, and the pre-specified Faculty-minus-Junior difference with a 95% confidence interval from the same Obuchowski-Rockette/DeLong reader-cluster bootstrap used for the primary endpoint. The ordered Junior-to-Senior-to-Faculty trend test is exploratory only. The Faculty–Junior contrast is interpreted under the two-one-sided-tests equivalence framework described in the Sample size subsection: the significance test evaluates the difference against zero, and the equivalence test evaluates it against the ±0.10 margin using the joint 90% confidence interval, with the resulting four-way reporting categorisation determining whether the result is described as a directional gradient, evidence against a clinically meaningful gradient, or inconclusive owing to power.

### 12. Correctness model and convergence fallback ladder

As a robustness analysis complementing the area-under-the-curve estimands, a generalised linear mixed model of trial-level correctness was specified as

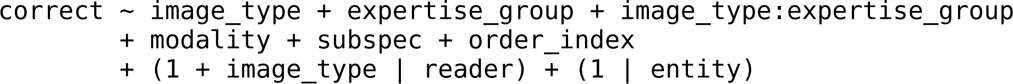

with image type taking the values authentic, Nano Banana 2, and gpt-image-2, and expertise group taking the values junior, senior, and faculty. The reported quantities are marginal predicted probabilities and odds ratios with 95% confidence intervals; areas under the curve are reported only from the Obuchowski-Rockette/DeLong analyses, not from this model. Because the full random-effects structure requires estimation of an unstructured reader-level covariance matrix under an unbalanced per-reader image-type allocation, singular-fit or non-convergence warnings were anticipated, and a fallback ladder was pre-specified in which the first specification returning a non-singular convergent fit becomes the reported model:

1. the full model above, with a correlated random slope for image type by reader;
2. a model in which the image-type random slope is collapsed to a binary AI-versus-authentic slope by reader; and
3. a model with random intercepts only for reader and for entity.

Convergence diagnostics (singular-fit warnings, parameter standard errors, log-likelihood, information criteria, and the condition number of the random-effects covariance) are reported, and if a reduced tier is used the Methods state which tier was reported and interpret the robustness conclusions under the constrained random-effects structure. The models were fitted with established mixed-model packages^23,24^.

### 13. Sensitivity analyses

The set of sensitivity analyses for the primary endpoint is capped, by pre-specification, at exactly four. First, a quality-restricted area under the curve was re-fitted on synthetic cells satisfying medical validity of 1, anatomical accuracy of at least 3, and image quality of at least 2, paired with the matched authentic counterpart of each retained entity; a supplementary variant that drops the image-quality criterion (retaining medical validity of 1 and anatomical accuracy of at least 3) is reported in parallel, because the calibration image-quality axis was the least reliable of the instrument, and a principal-component factor-score variant of the composite quality covariate is reported as a construct-validity check. Second, the mixed model was re-fitted excluding the quality-assurance faculty readers, which reduces the Faculty stratum and widens the minimum detectable difference; this sensitivity is interpretive (gradient-direction robustness) rather than confirmatory, and faculty-specific estimates are reported descriptively if the residual Faculty count falls below 12. Third, the model was re-fitted excluding readers with fewer than 14 days of washout between quality assurance and the reader task; because all quality-assurance faculty met the 14-day preferred threshold at the broadcast date, this sensitivity is vacuous by design at broadcast and is reported as hardened by full compliance with the preferred washout threshold.

Fourth, separate models were fitted for cells passing and failing the principal-investigator visual-realism assessment, as a descriptive comparison rather than a hypothesis test. A realism finding is reported only if the primary pool and the quality-restricted analysis point in the same direction; consistent with the primary-analysis subsection, no deployable-classifier claim is made from the human-reader area under the curve, which can at most identify provenance screening as a hypothesis for dedicated classifier development and external validation.

Beyond these four, the per-model coverage and missing-not-at-random analyses for the vendor-refused and rule-dropped cells, the abstention-proxy and completion-cutoff handling, and the paired-loss handling are reported as supplementary sensitivity reports of the missingness structure and do not count against the four-entry scope cap; the response-duration audit, the aspect-ratio descriptive analysis, and the high-completion subgroup are descriptive only and are not promoted to sensitivity status.

### 14. Exploratory analyses

The cue-category distribution by expertise stratum (without an accuracy join) is reported as a descriptive contingency table with a nominal χ² test, and an optional natural-language taxonomy of the free-text reasoning is reported as exploratory only. A discrete pre-launch taxonomy-validation pilot was not conducted as a separate step; instead, the adequacy of the seven prespecified cue categories is assessed at analysis from the trial-level cue distribution and from the rate at which readers selected the “guessed or uncertain” category or supplied free-text reasoning not captured by the seven categories, and any resulting limitation of the taxonomy is reported descriptively. The prespecified cue-by-accuracy analysis cross-tabulates the seven-category cue against trial-level accuracy within each of the three expertise strata, reporting per-stratum 7-by-2 tables, χ² tests with continuity correction, Cramér’s V with a 1,000-iteration bootstrap 95% CI at seed 42, per-cue accuracy proportions with Wilson 95% CIs, and a Fisher exact substitute where any expected cell count falls below five. Cramér’s V is the headline effect-size metric within each stratum; the per-cue Wilson confidence intervals are reported as descriptive disaggregation rather than as independent hypothesis tests. Although this analysis is reported as a nominal-confidence-interval descriptor rather than as a family-wise-error seat, a Benjamini–Hochberg false-discovery-rate supplement across the stratum-wise tests and the per-cue proportion tests is provided so that readers may apply false-discovery-controlled interpretation; the false-discovery-adjusted values are supplementary and do not modify the nominal inference. Prespecified subgroup analyses by reader subspecialty match and by board-certification recency, and an image-quality-by-expertise interaction, are reported as exploratory and descriptive. Patient-demographic fairness (accuracy stratified by patient sex, age, or race)v cannot be assessed because the authentic arm is sourced from a teaching repository for which per-image patient demographics are not consistently available; this constraint is reported as a limitation, paired with the corresponding fairness-reporting item, and no retrospective demographic imputation is attempted.

### 15. Multiplicity

Multiplicity is controlled by a fixed-sequence hierarchical gatekeeping rule. The primary endpoint (H1) is tested at two-sided α = 0.05; the Faculty–Junior contrast (H2) is tested at two-sided α = 0.05 gated on the significance of H1; and the contrast between the two synthetic arms is reported with a nominal p-value and a 95% confidence interval, labelled confirmatory only if H2 is also significant. The quality-assurance reliability statistics (Krippendorff’s α across the six axes and Fleiss’ κ across the artefact categories) are reported descriptively against reliability tiers without a hypothesis-test α, with a descriptive step-down correction retained for the artefact-category family and the prior hard reliability gates removed. The prespecified cue-by-accuracy analysis is reported as a nominal-confidence-interval descriptor within each stratum and is not subject to multiplicity correction beyond the hierarchical gatekeeping; to limit the inferential surface of this analysis, its primary inferential currency is the three per-stratum Cramér’s V values (each with a bootstrap 95% confidence interval) rather than the per-cue proportions, the per-cue Wilson intervals are reported as descriptive disaggregation rather than as independent hypothesis tests, and a Benjamini–Hochberg false-discovery-rate supplement is provided for readers who prefer family-wise-controlled interpretation. All other secondary analyses are reported with nominal 95% confidence intervals as the primary inferential currency, with the exploratory analyses carrying no multiplicity correction. The cumulative count of formal hypothesis tests is small (on the order of the primary endpoint, the key secondary contrast, and a prevalence-gated artefact-reliability family), and no formal multiplicity correction beyond the hierarchical gatekeeping is operative; descriptive reliability-tier reporting follows recognised practice in the multi-reader reliability literature^19,20^.

### 16. Aspect-ratio cue

A residual extra-diagnostic provenance cue persists in the pool: the synthetic arms are produced in near-square aspect ratios, whereas authentic source images vary in aspect ratio, and this difference is not removed by the crop-free normalisation, which prioritises anatomy preservation over distribution matching. Image-format harmonisation equalised resolution, file size, compression, filename, and metadata across the three arms (see the Image-format harmonisation subsection), so native aspect ratio is the principal residual format attribute that was not matched; the harmonisation surface and its single residual are therefore bounded and disclosed rather than open-ended. The cue is documented in three ways: a per-arm aspect-ratio distribution is reported as descriptive quality control; the pooled area under the curve is reported descriptively, stratified by aspect-ratio quartile together with each quartile’s arm composition, so that the magnitude of any aspect-ratio-driven contribution to the primary endpoint is visible; and the cue is acknowledged in the Limitations as a residual confounder that the normalisation chose not to mask, because anatomy distortion would compromise diagnostic content and symmetric padding would introduce artificial borders while leaving the content-aspect cue intact. In addition, the separability of the arms by native aspect ratio alone is reported as a descriptive ceiling — the area under the curve of a univariate classifier on aspect ratio computed from the pool manifest, not from reader responses — so that the maximum contribution the format cue could make to the reader endpoint is bounded; and within aspect-ratio strata in which the synthetic and authentic arms overlap, the reader area under the curve cannot be produced by the format cue and is read as the de-confounded estimate. The aspect-ratio cue is reported as a stratification axis, a bounded separability ceiling, and a transparent limitation, not as a covariate-controlled estimate, and is not promoted to a sensitivity analysis under the four-entry scope cap. For transparency of the authentic arm, the 19 view/specification alignment refinements applied during the manifest lineage are conservatively classified as subjective view-preference refinements (18 retain the same source case and the single case with a changed source URL lacks a per-swap objective normalisation-failure criterion) and are described in prose as view or specification alignment refinements rather than as corrections of incorrect images; the classification is for reviewer transparency and is not an eligibility gate.

### 17. Reporting framework

The study is reported against the AI-validation reporting frameworks appropriate to its components, each cited with its base guideline where one exists. STARD-AI^25^ alongside the base STARD 2015 statement^26^ applies by analogy: the reader task is analyzed with diagnostic-accuracy (area-under-the-curve) methods, with image provenance serving as a deterministically known reference standard rather than a clinical target condition requiring adjudication. The 2024 CLAIM update^27^ alongside the base CLAIM statement^28^ applies to the AI-image-generation pipeline. MI-CLEAR-LLM^29^ applies to the S5 large-language-model-as-a-judge content validator in the upstream teaching-content pipeline. Reporting of that same S5 component additionally follows the LLM-methods items of the TRIPOD-LLM statement^30^; items specific to LLM prognostic or diagnostic models and to clinical deployment are not applicable, because the large language model is used as a content-quality validator rather than as a patient-facing prediction model. Per-framework item-level compliance is reported in the Supplement, with each item flagged as present, partial, missing, or not applicable against its manuscript location; checklist totals are recomputed from the per-item matrix at build time rather than hand-tallied.

### 18. AI-use disclosure

The authors used the following generative AI tools during the conduct, analysis, and reporting of this study, each reported with vendor, version identifier, access channel, date range of use, responsible party, and intended task, consistent with the disclosure requirements of MI-CLEAR-LLM^29^ and the LLM-methods items of TRIPOD-LLM^30^. The Nano Banana 2 model (Google DeepMind; Gemini 3.1 Flash Image Preview) was accessed through the Google Generative AI SDK between 2026-04-08 and 2026-04-29 (frozen at frame lock), with the first author responsible, to generate the Nano Banana 2 synthetic arm; gpt-image-2 (OpenAI) was accessed through the OpenAI Batch API over the same window, with the first author responsible, to generate the gpt-image-2 synthetic arm. These synthetic radiologic images are the object of study and were presented to readers as a synthetic arm; they were not used to illustrate the manuscript. Claude (Anthropic; versions 3.7 Sonnet, Opus 4.7, and Opus 4.8) was used between 2025-12-28 and 2026-06-06, with the first author responsible, for protocol drafting and revision support, reporting-compliance auditing, citation cross-checking against PubMed and CrossRef, and language editing; all outputs were reviewed and verified against primary sources by the authors. Consistent with the companion Paper 1 disclosure, Claude served as a manuscript-preparation and analysis-support assistant and was not part of the validated content-generation pipeline. The upstream S5 LLM-as-a-judge content validation of the entity-table and card stages was performed by a separate generative model, documented in the AI-mediated prompt-refinement subsection of the Methods and in Supplementary Methods S3 rather than attributed here.

ChatGPT (OpenAI) was accessed through its web interface during the pre-submission review cycle, with the first author responsible, to cross-check statistical, reporting, and arithmetic claims as part of a multi-stage internal review; it was not used to generate any endpoint estimate, to determine reader eligibility, or to perform statistical inference.

Generative AI was not used to create, modify, or curate any image presented to readers beyond the two synthetic arms whose generation is the object of study; to compute, estimate, or interpret any endpoint; to determine reader eligibility, stratification, randomisation, or assignment; or to perform statistical inference, all of which were executed in R and Python with the documented packages and seeds. Any anatomical icons or illustrative non-data figures in the manuscript are modified from SMART Servier Medical Art^31^ under a Creative Commons Attribution 4.0 International licence. The target journal permits AI-generated content with disclosure; the synthetic radiologic images are evaluated as the object of investigation rather than as figures within the meaning of the journal’s AI-image policy, and their disclosure as study material is documented with vendor, version, freeze date, and per-image provenance. The generative models used to create the synthetic arms are commercial products (Google DeepMind and OpenAI), accessed under standard paid application-programming-interface terms; the authors have no financial relationship with, or commercial interest in, either vendor, and no vendor had any role in study design, data analysis, or the decision to publish.

### 19. Pre-registration, data, and code availability

The study was preregistered at the Open Science Framework (registration of record https://osf.io/zrwy8; DOI 10.17605/OSF.IO/ZRWY8;^16^), filed and made public on 2026-06-04 KST before the reader task was broadcast; the locked statistical analysis plan is archived with the registration. The preregistration deviation log records the fixation of the seven-category cue item, the prespecification of the cue-by-accuracy secondary analysis on 2026-05-23, and the confirmation of free-text cue capture, all lodged before broadcast. De-identified reader-level response data, normalised-image hashes, and the assignment matrix will be released on Zenodo under a Creative Commons Attribution 4.0 International licence at publication, with source images released as a metadata catalogue (authentic-arm case identifiers and URLs; synthetic-arm prompts and image hashes) subject to source-repository attribution policy. The R and Python analysis scripts are archived on Zenodo with a DOI assigned at publication; the primary correctness model, the quality-restricted variants, the cue-by-accuracy analysis, the alternative-rule audit, and the principal-component construct-validity refit are committed as named scripts.

## 20. Deviations and amendment policy

After the pre-dispatch lock (locked before any reader data collection), no further change to the primary-pool composition, the drop rule, or the reader-exclusion criteria was made except for documented technical deployment failures occurring before reader exposure; the lock governs the pool composition, the consensus-fail drop rule, the reader-exclusion criteria, the four-entry sensitivity scope cap, and the reliability reporting regime. The lock does not govern measurement-instrument analysis prespecifications added before any reader data collection, such as the cue-by-accuracy secondary analysis, the principal-component construct-validity refit, the image-quality-dropped quality-restricted variant, and the false-discovery-rate supplement, all prespecified before broadcast. Any issue arising after dispatch is handled as a protocol deviation, a descriptive note, or one of the four prespecified sensitivity analyses, never as a primary-pool reconstruction, and any post-dispatch deviation is recorded in a dated deviation log with an explicit statement of whether it affected primary-endpoint estimation. The locked analysis plan is the single canonical statistical analysis plan; earlier amendment documents remain as the audit trail. Result-direction interpretations were precommitted before dispatch to four direction-specific framings (near-chance discrimination, modest-but-insufficient discrimination, no Faculty advantage, and Faculty advantage present), each to be reported according to the observed result so as to constrain narrative drift between draft and submission.

Participant-list reconciliation is documented at four levels: the institutional review-board roster; the raw dispatch list after excluding acknowledgement-only named readers and reconciling a registration-only invitee; the final dispatch set of 60 readers after excluding the co-investigator who developed the pipeline to preserve evaluator–investigator independence (reducing the matrix from a 61-reader, 3,660-row baseline to 60 readers and 3,600 rows); and the completed analysis cohort, reported at analysis time. Historical registration responses were never edited; the reconciliation record is the sole audit source, and this is participant-list reconciliation rather than an outcome-related design change. Because the pre-dispatch lock was followed by a cluster of prospectively documented adjustments before any reader exposure — two rater-confirmed clerical corrections, the five-cell consensus-fail drop, the image-format harmonisation, the view/specification swap labelling, the participant reconciliation, and the cue-by-accuracy prespecification — these are disclosed collectively in a single Abstract sentence, an enumerated Limitations paragraph, and Supplementary Table S1 (the post-lock timeline), which records each adjustment with its date, authorising clause, and artefact hash, together with the broadcast-date lineage (the broadcast date was moved forward, before data collection, so that all quality-assurance faculty met the 14-day preferred washout threshold). Item-level compliance with the MI-CLAIM reporting checklist and a summary of reader diversity and representativeness are provided in Supplementary Tables S2 and S3, respectively.

### 21. Manuscript framing rule

The manuscript frames the study in terms of synthetic-image provenance, AI literacy, expert-supervised training, and provenance-discrimination ability, and avoids the language of fraud or deception. A null or near-chance Faculty result is a publishable finding that shifts the framing toward the conclusion that even experts have limited provenance-discrimination ability, supporting the development and prospective evaluation of systematic safeguards and AI-literacy curricula. The pre-specified intended use of the study is to inform radiology AI-literacy curricula and image-provenance training — specifically, whether expert-supervised AI-literacy training is necessary and whether transferable provenance cues exist for teaching. Secondary implications, including editorial figure-provenance auditing, journal anti-fabrication screening, and educational-image curation, are exploratory hypotheses rather than validated uses: reaching an area under the curve of at least 0.75 with a lower confidence bound of at least 0.70, jointly satisfied by the primary pool and the quality-restricted concordance criterion, would identify these as directions warranting dedicated classifier development and external clinical-spectrum validation, and none is licensed by the human-reader area under the curve alone. The study does not claim to validate any deployed image-provenance classifier; a demonstration of curriculum effectiveness is delegated to a companion study in radiology medical education from our group^17^.

The reliability gate (α ≥ 0.50) is not reported as establishing reliability; the manuscript reports the α point estimate and confidence interval against the reliability tiers and uses the tier system, not the operational gate, for substantive interpretation.

## Results

All 60 invited readers completed the visual Turing test and constitute the full analysis set (Faculty, 20; Senior, 18; Junior, 22). The locked pool comprised 241 displayable cells across 82 entities, nine subspecialties, and six modalities (60 readers x 60 trials = 3,600 reader–image observations; study flow, Figure 1). Baseline reader characteristics are shown in Table 1.

**Figure 1.**
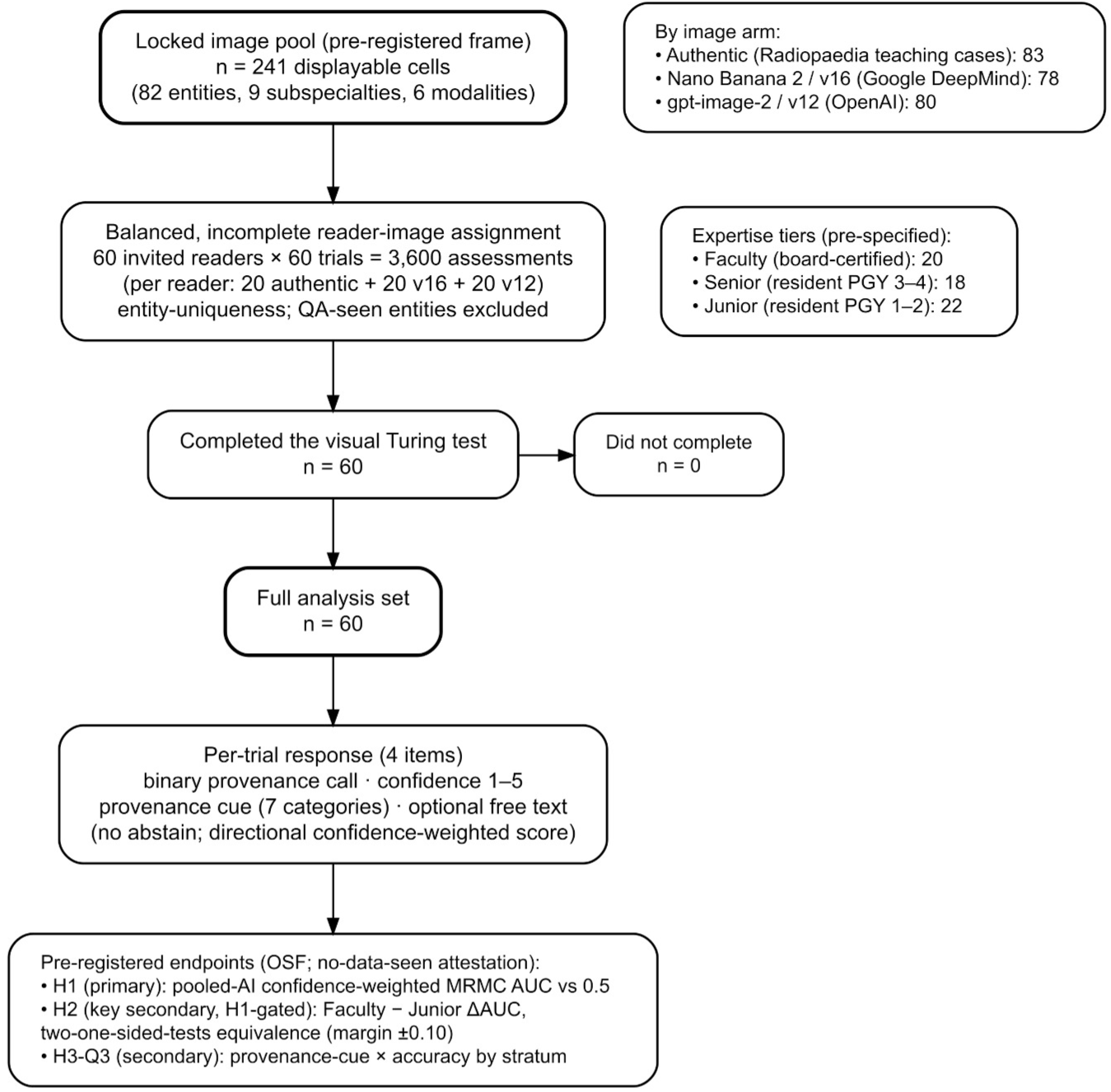
Study design and reader flow. The pre-registered, locked image pool comprised 241 displayable cells (82 distinct entities spanning nine subspecialties and six modalities: computed tomography, magnetic resonance imaging, ultrasonography, radiography, mammography, and angiography), divided into three arms: 83 authentic teaching-repository cases and 158 AI-generated images from two contemporary commercial generators (78 from Nano Banana 2 [Google DeepMind]; 80 from gpt-image-2 [OpenAI]). Sixty invited readers each judged 60 trials under a balanced, incomplete reader–image assignment (20 authentic, 20 Nano Banana 2, and 20 gpt-image-2 per reader; 60 × 60 = 3,600 reader–image observations), with entity-uniqueness enforced and quality-assurance–seen entities excluded. Readers were pre-stratified into three expertise tiers (Faculty, board-certified, n = 20; Senior, residents in postgraduate years 3–4, n = 18; Junior, residents in postgraduate years 1–2, n = 22). All 60 readers completed the test (full analysis set, n = 60). Each trial recorded a binary provenance call, a confidence rating (1–5), one of seven provenance-cue categories, and optional free text. Pre-registered endpoints (Open Science Framework, no-data-seen attestation) were H1 (pooled-AI confidence-weighted multi-reader multi-case area under the curve versus 0.5), H2 (H1-gated Faculty-minus-Junior difference in the area under the curve under a two one-sided tests equivalence framework, margin ±0.10), and H3-Q3 (provenance-cue category × trial-level accuracy by stratum).

**Table 1.** Reader characteristics by expertise tier.

| Characteristic | Junior (n=22) | Senior (n=18) | Faculty (n=20) | Total (n=60) |
| --- | --- | --- | --- | --- |
| Board-certified, n (%) | 0 (0) | 0 (0) | 20 (100) | 20 (33) |
| Postgraduate-year band | PGY 1–2 | PGY 3–4 | board-certified | — |
| Years since board certification, median (IQR) | not applicable (in training) | not applicable (in training) | 6 (4–7)* | 6 (4–7)* |
| Declared subspecialty, n | in training | in training | general radiology 10, musculoskeletal 5, interventional 2, breast 1, cardiovascular 1, neuro/head-and-neck 1 | — |
| Also served as a QA rater, n | 0 | 0 | 6 | 6 |
| Washout days at broadcast | — | — | all $\geq 14$ | all $\geq 14$ |
| Completed all 60 trials, n (%) | 22 (100) | 18 (100) | 20 (100) | 60 (100) |

\* Years since board certification was reported by 12 of 20 faculty on the enrollment form. Subspecialty was collected for board-certified faculty only (those who did not declare one are recorded as general radiology); residents are in training and were not assigned a subspecialty. Reader sex and age were not collected. Image subspecialties (the nine radiologic categories spanning the pool) are distinct from reader subspecialty.

Readers discriminated AI-generated from authentic images modestly above chance. The pooled confidence-weighted area under the curve was 0.71 (95% CI, 0.69 to 0.74), estimated as the reader-averaged area under the curve conditional on the locked 241-image pool, with a reader-cluster bootstrap confidence interval; the 95% confidence interval excluded the null value of 0.5. This estimate falls in the pre-specified modest tier (0.60 to 0.75), in which discrimination is real but insufficient to serve as a standalone provenance filter (Figure 2). A supplementary DeLong interval (0.71; 95% CI, 0.69 to 0.73) and a reader-and-case generalization bootstrap (0.71; 95% CI, 0.68 to 0.75) were concordant; inference beyond the pool rests on this case-inclusive interval. The two commercial generators were similar in detectability (Nano Banana 2, 0.70; gpt-image-2, 0.73; absolute difference, 0.02), below the pre-specified model-stratified pivot of 0.10, so the pooled estimate is reported as the headline (Table 2).

**Figure 2.**
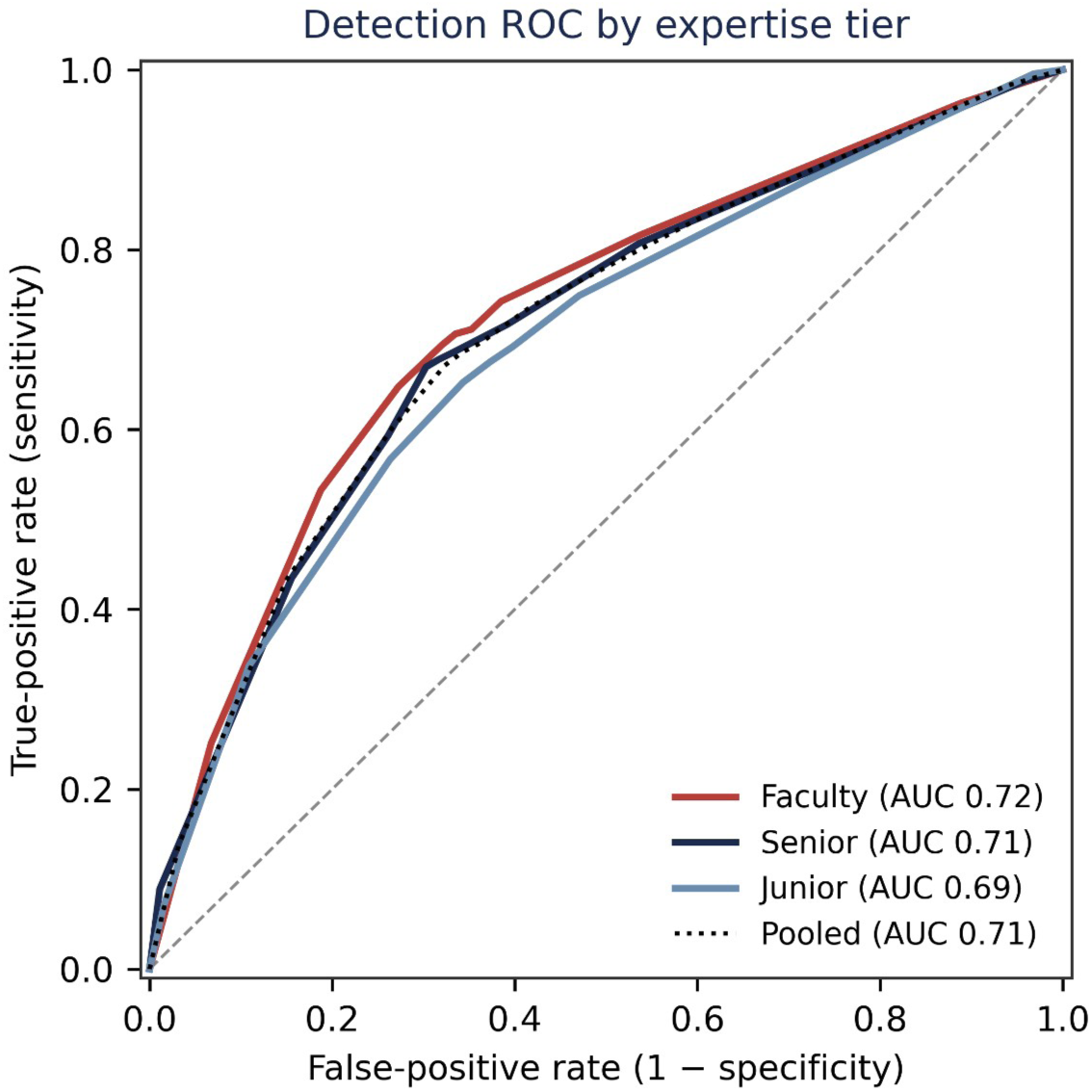
Detection ROC by expertise tier. Confidence-weighted receiver-operating-characteristic curves for discriminating AI-generated from authentic radiologic images, plotted for each expertise tier and for the pooled cohort. The areas shown in the figure are the empirical curve areas computed from the confidence-weighted provenance-suspicion score across all reader–image observations within each stratum (Faculty 0.72, Senior 0.71, Junior 0.69, pooled 0.71); these differ marginally from the pre-specified confidence-weighted, reader-averaged multi-reader multi-case area-under-the-curve estimates reported in Table 2 (Faculty 0.73, Senior 0.72, Junior 0.69, pooled 0.71), which are the inferential estimates for all hypothesis tests. The diagonal dashed line denotes chance (area under the curve 0.5).

**Table 2.** Primary detection performance — confidence-weighted reader-averaged MRMC AUC (H1) and expertise contrast (H2)

| Comparison | AUC | DeLong 95% CI | Reader-cluster boot 95% CI | Tier | Deployable† |
| --- | --- | --- | --- | --- | --- |
| **Pooled AI vs authentic (H1)** | 0.71 | 0.69 to 0.73 | 0.69 to 0.74 | modest | No |
| Nano Banana 2 vs authentic | 0.70 | — | 0.67 to 0.73 | modest | No |
| gpt-image-2 vs authentic | 0.73 | — | 0.70 to 0.75 | modest | No |
| ΔAUC (gpt-image-2 – Nano Banana 2) | 0.023 | — | 0.00 to 0.05 (no pivot; <0.10) | — | — |

**H2 expertise contrast (TOST, margin $\pm 0.10$ ):** Faculty AUC 0.73 vs Junior AUC 0.69; $\Delta$ AUC(F–J) = 0.04, 95% CI –0.02 to 0.10 (includes 0; no significant gradient); TOST 90% CI –0.01 to 0.09, **equivalence established**. H1 gate passed (H1 CI excluded 0.5). Monotonic point-estimate trend: Junior 0.69 < Senior 0.72 < Faculty 0.73 (descriptive). Reader-and-case generalization bootstrap: 0.71 (0.68 to 0.75).
† Deployable-classifier admissibility: AUC $\geq 0.75$ and lower 95% CI $\geq 0.70$ , jointly with the quality-restricted sensitivity analysis (Supplementary Table S4); not met at any stratum. AUC tiers: near-chance 0.50–0.60, modest 0.60–0.75, clinically meaningful $\geq 0.75$ .

Point estimates ordered with expertise (Junior, 0.69 [95% CI, 0.65 to 0.74]; Senior, 0.72 [95% CI, 0.67 to 0.76]; Faculty, 0.73 [95% CI, 0.69 to 0.78]), but the pre-specified Faculty-minus-Junior difference in the area under the curve was 0.04 (95% CI, −0.02 to 0.10), and the 95% confidence interval included zero; a significant expertise gradient was therefore not established. Under the two one-sided tests at the +/−0.10 equivalence margin, the joint 90% confidence interval (−0.01 to 0.09) lay within the margin, establishing equivalence. No stratum met the pre-specified standalone-classifier tier (an area under the curve of at least 0.75 with a lower 95% confidence bound of at least 0.70); the Faculty point estimate (0.73, lower bound 0.69) remained in the modest tier.

The reader-selected provenance-cue category was associated with trial-level accuracy within every expertise stratum (Faculty Cramér’s V = 0.15, 95% CI 0.11 to 0.21, P < 0.001; Senior 0.14, P = 0.002; Junior 0.14, P < 0.001), indicating that provenance-cue informativeness varied by category. Per-stratum estimates are reported in Table 3.

**Table 3.** Provenance-cue category × trial-level accuracy, by expertise stratum (H3)

| Stratum | $\chi^2$ (df=6) | p | Cramér's V<br>[95% CI] | Most accurate<br>cue (accuracy) |
| --- | --- | --- | --- | --- |
| Junior | 26.78 | $1.6 \times 10^{-4}$ | 0.14 [0.11, 0.21] | pathology<br>realism (0.81) |
| Senior | 20.71 | $2.1 \times 10^{-3}$ | 0.14 [0.11, 0.21] | pathology<br>realism (0.76) |
| Faculty | 25.68 | $2.6 \times 10^{-4}$ | 0.15 [0.11, 0.21] | image-quality<br>artifact (0.77) |

$\chi^2$ with Yates continuity correction; Cramér's V with a 1,000-iteration bootstrap 95% CI (seed 42); per-cue Wilson 95% CIs in the Supplementary Appendix; Benjamini–Hochberg FDR control. Pre-specified before data lock.

The primary finding was robust across every pre-specified sensitivity analysis (Supplementary Table S4): a quality-restricted area under the curve limited to high-rated synthetic images (0.69), restriction to complete-coverage entities (0.72), and exclusion of quality-assurance–rater readers (0.71) all remained in the modest tier with no divergence beyond the pre-specified thresholds, and none reached the deployable-classifier threshold. Higher quality-assurance–rated synthetic image quality was associated with lower reader detection (odds ratio, 0.84; 95% CI, 0.80 to 0.89; Supplementary Table S5); because the graded quality axes had limited inter-rater reliability (Supplementary Table S6), this realism gradient is reported as an exploratory, covariate-level association that does not bear on the primary endpoint. Detection varied across modalities (Figure 3 and Supplementary Table S7), from 0.63 (radiography) to 0.79 (ultrasound, the only modality reaching the deployable tier), but these estimates are descriptive, not powered for stratified inference. In post-hoc exploratory analyses, reader confidence tracked accuracy (accuracy rising from 0.55 at the lowest confidence level to 0.83 at the highest; Supplementary Table S8), yet 27.5% of high-confidence judgments were incorrect (Figure 4); per-reader discrimination varied widely (range, 0.45 to 0.94), with three readers performing at or below chance. Representative synthetic images spanning the realism tiers and the cells excluded at the quality-assurance gate are shown in Figure 5 (Panels A and B). The individual images most frequently misclassified by readers, all AI-generated, are shown in Panel C (post-hoc exploratory): the most-misclassified image, a gpt-image-2 pediatric radiograph, was misjudged by 91% of the readers who evaluated it. In a post-hoc descriptive comparison (not a pre-specified sensitivity analysis), the unweighted binary-call area under the curve was 0.67, lower than the pre-registered confidence-weighted estimate of 0.71, indicating that the confidence weighting contributed modestly and that discrimination remained within the modest tier without it.

**Figure 3.**
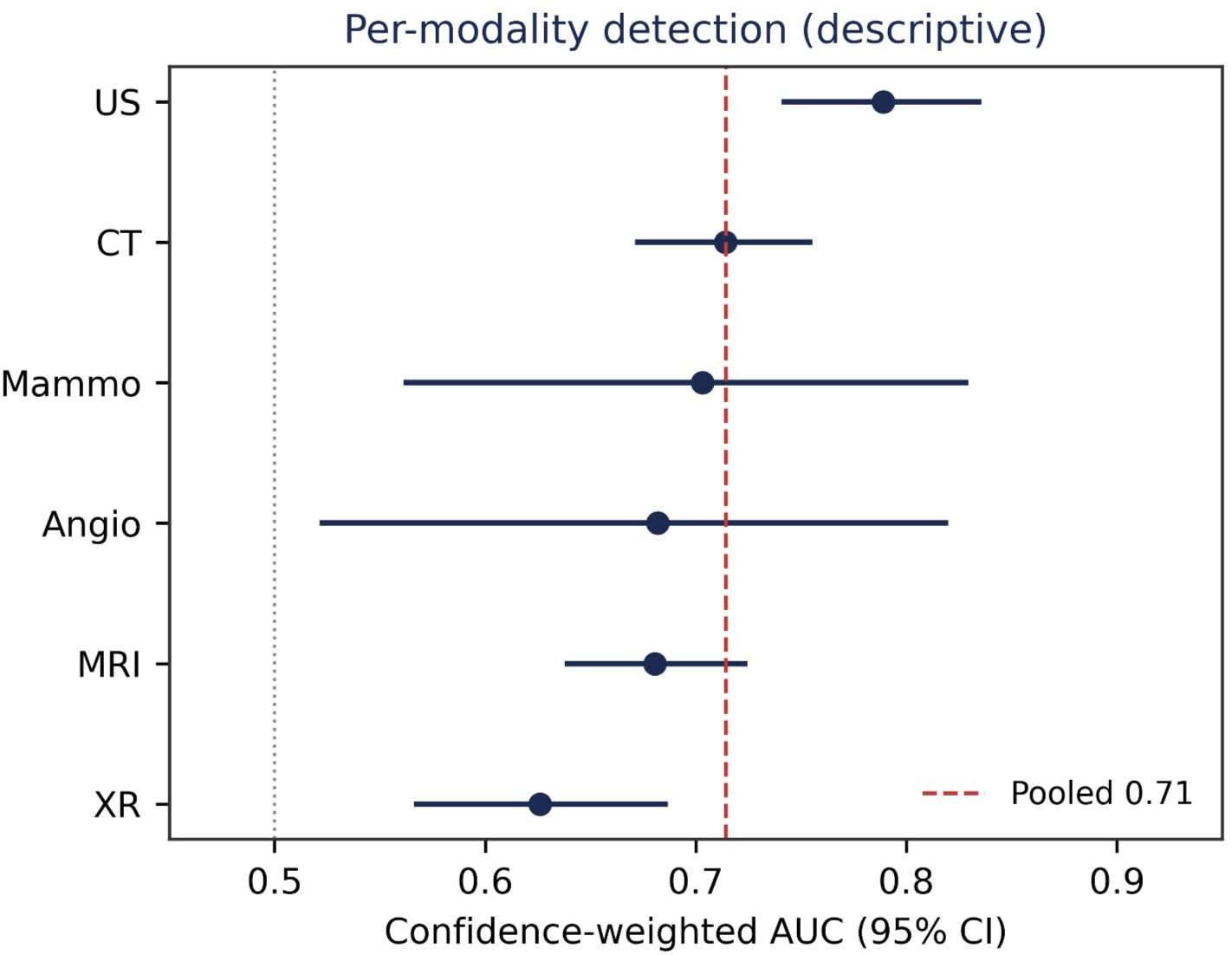
Per-modality detection (descriptive). Forest plot of the confidence-weighted area under the curve with 95% confidence interval for each of the six modalities, ordered by point estimate. Detection ranged from 0.63 (radiography) to 0.79 (ultrasonography, the only modality reaching the deployable tier). The coral dashed line marks the pooled estimate (0.71) and the dotted line marks chance (0.5). These per-modality estimates are descriptive and were not powered for stratified inference.

**Figure 4.**
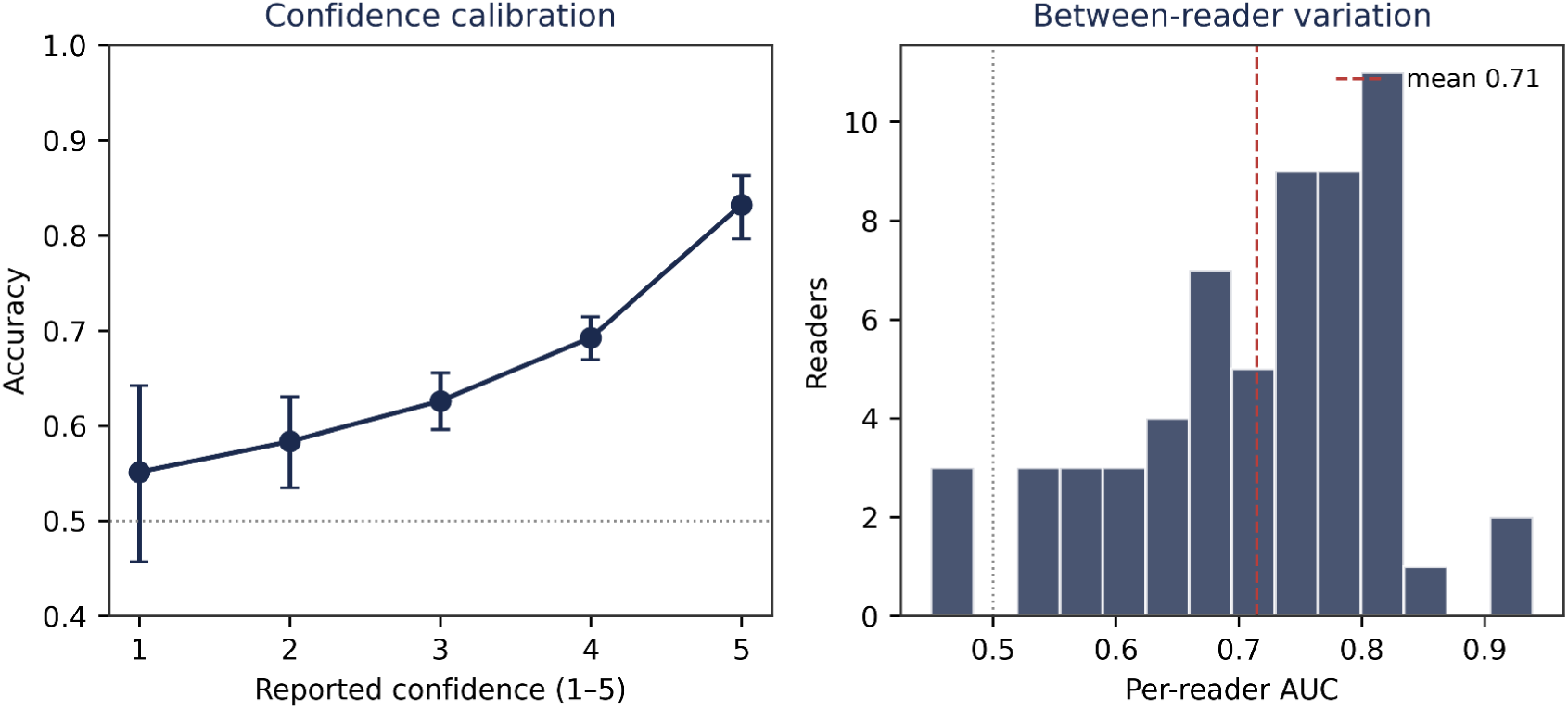
Confidence calibration and between-reader variation. Left panel: trial-level accuracy by reported confidence (1–5) with Wilson 95% confidence intervals; accuracy rose monotonically from 0.55 at the lowest confidence level to 0.83 at the highest (dotted line, chance), yet a substantial fraction of high-confidence judgments were incorrect. Right panel: distribution of per-reader area under the curve across the 60 readers (range, 0.45 to 0.94; coral dashed line, mean 0.71), with three readers performing at or below chance, indicating wide between-reader variation in discrimination ability.

**Figure 5.**
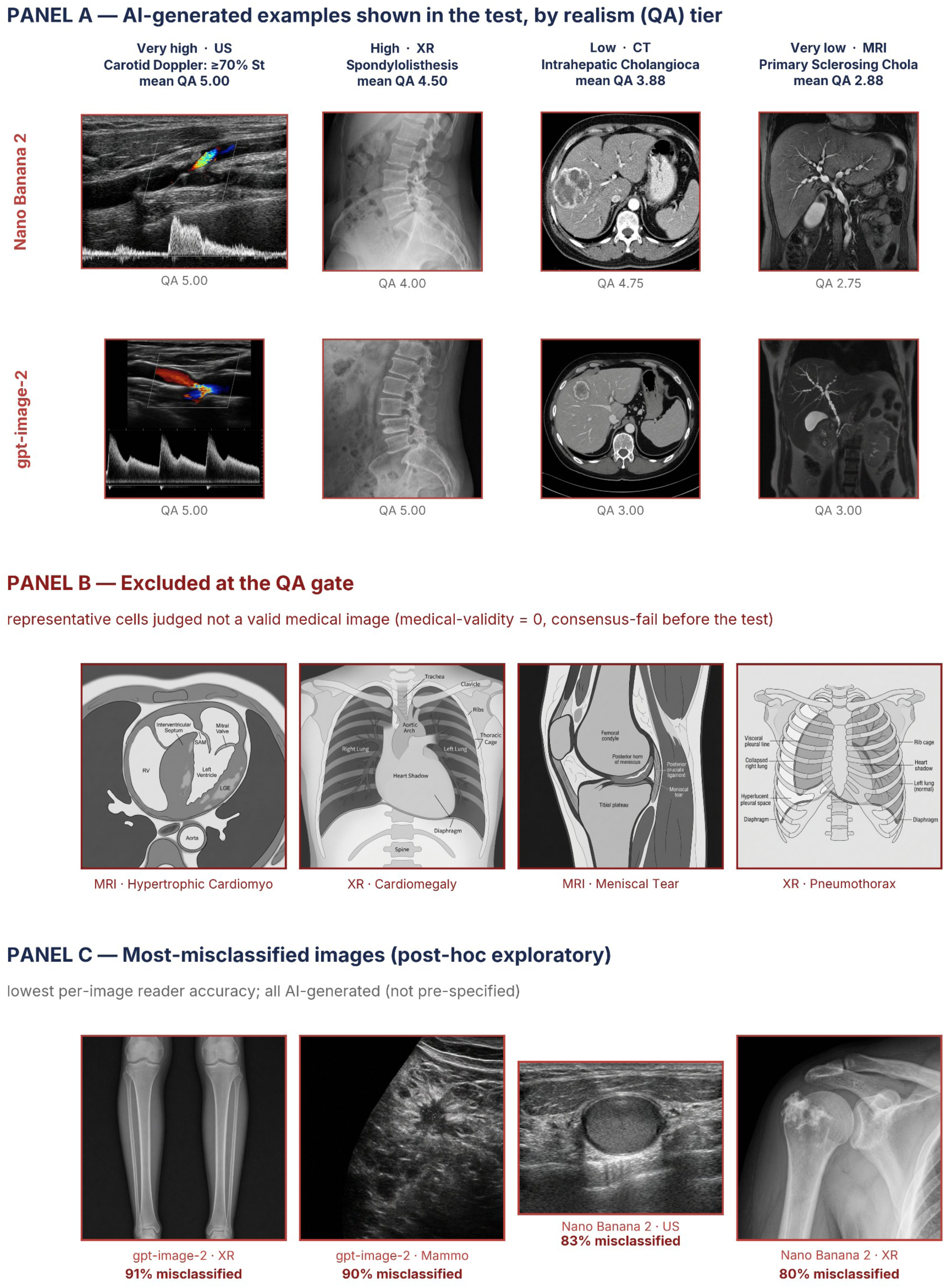
AI-generated images evaluated in the test: matched examples by realism tier, cells excluded at the quality-assurance gate, and the most-misclassified images. Panel A (matched examples by realism tier): each column is one entity, ordered from highest to lowest synthetic-quality tier left to right (QA score = mean of the four 1–5 quality-assurance axes); the two rows show the matched Nano Banana 2 and gpt-image-2 outputs for that entity, spanning four modalities. Panel B (excluded at the quality-assurance gate): representative cells removed before the test because judged not valid medical images (medical-validity = 0, consensus-fail). Panel C (most-misclassified images; post-hoc exploratory, not pre-specified): the individual images with the lowest per-image reader accuracy, annotated with the percentage of readers who misclassified each; all are AI-generated, and the worst (a gpt-image-2 pediatric radiograph) was misjudged by 91% of the readers who evaluated it. Images are the format-harmonized versions shown to readers; native aspect ratio and colour are preserved. Only the author-generated AI arms are reproduced; the authentic comparator was drawn from the Radiopaedia teaching repository (CC BY-NC-SA 3.0), and its case identifiers and URLs will be provided in the data catalogue released at publication, rather than reproduced here. Aspect-ratio sensitivity analyses are summarized in Supplementary Table S9.

## Discussion

In this pre-registered, multimodal visual Turing test, 60 invited radiology faculty and trainees judged authentic and frontier-generated radiologic images across six modalities and nine subspecialties. The pooled confidence-weighted multi-reader multi-case area under the curve, conditional on the locked image pool, was 0.71 (95% CI, 0.69 to 0.74), above chance but within the pre-specified modest tier (0.60 to 0.75), where discrimination is real yet insufficient as a standalone filter. The Faculty-minus-Junior contrast was 0.04 (95% CI, −0.02 to 0.10), including zero, and the two one-sided tests established equivalence within the pre-specified +/−0.10 margin, so any expertise gradient is bounded within this margin, although the study was not powered to exclude a smaller (0.05 to 0.10) advantage. No stratum reached a dependable safeguard: even board-certified faculty (0.73) did not meet the pre-specified standalone-classifier tier, and the finding held across every sensitivity analysis.

These findings confirm and extend the single-generator, single-modality chest-radiograph results of Tordjman and colleagues^1^, who reported that only a minority of radiologists spontaneously recognised generated radiographs, that years of experience were unrelated to detection, and that inter-reader agreement was fair (Fleiss κ = 0.31). We reproduce the central lesson (frontier-generated radiologic images are difficult to distinguish from authentic ones; detection is graded, not categorical) while broadening it along three axes prior work left open: from radiography alone to cross-sectional and ultrasound modalities central to subspecialty practice; from one generator to two contemporary commercial generators similar in detectability (absolute difference, 0.02, below the 0.10 model-stratified pivot), so neither afforded a decisive advantage; and from anecdotal cue description to a prespecified seven-category cue instrument linked to accuracy. Inter-reader concordance was consistent with the prior fair agreement: per-reader discrimination varied widely (Figure 4), indicating that limited concordance is intrinsic to the task, not a measurement defect. And consistent with their observation that years of experience were unrelated to detection, our pre-registered training-tier contrast showed no significant difference (equivalence within +/−0.10); the modest point-estimate ordering is interpreted cautiously given potential confounding by subspecialty familiarity and case mix.

The clinical relevance lies in an escalating threat to image-record integrity. Concern began with manual manipulation and duplication of genuine images, detectable when at all by trained human scrutiny^4^; it advanced to artificial-intelligence alteration of genuine images, as when a published clinical-image case was retracted after an AI tool altered the figure, caught only by post-publication scrutiny^5,32^. The present study addresses the more severe end of this arc (full generation rather than partial alteration) and asks whether unaided expert review is a dependable defence as generators improve^6,7^; two contemporary systems already sit at a level where it is not. The authentic comparator here was drawn from a teaching repository, so whether these findings extend to consecutive clinical-acquisition images, which differ in spectrum and quality, remains untested.

The images evaluated here were produced by the upstream pipeline our group developed for radiology teaching-material synthesis^18^ and were withheld from educational distribution out of the perceptual-harm concern motivating this work, which sits between that pipeline and our educational follow-up^17^. Because unaided expert review was an inadequate standalone safeguard, the principal implication is for radiology AI-literacy curricula. Unverified AI-generated images increasingly circulate on social-media platforms as informal learning material, risking distortion of the perceptual reference trainees calibrate against. Our findings favour, for image-based learning, sources documenting each case’s origin (peer-reviewed literature, textbooks, moderated teaching repositories) over collections without a provenance trail. For governance, the findings motivate a layered approach rather than reader vigilance: documented source provenance where feasible, cryptographic content provenance, watermarking, and declaration of image processing. We frame this as a consideration, not a recommendation: acquisition metadata is strippable and spoofable, and we evaluated no provenance intervention.

This study has several limitations. First, the synthetic arm used two commercial generators frozen at fixed snapshots and does not sample the full space of systems; the escalation contrast illustrates a trajectory, not an exhaustive survey. Second, the endpoint is perceptual distinguishability under reader conditions, not deployment-validated automated classification; external evaluation on independent cohorts would be required for any provenance-screening claim. Third, the authentic arm came from a public teaching repository, so scanner, acquisition, and demographic details are inconsistently available, and the estimand is scoped to teaching-quality authentic images consistent with the AI-literacy intended use. Fourth, native image aspect ratio partially distinguishes the arms, because crop-free normalisation preserved anatomy rather than matching aspect-ratio distributions. A univariate aspect-ratio classifier reached an area under the curve of 0.70; reader discrimination (0.71) exceeded this format ceiling by only a narrow margin (0.71 versus 0.70), and within the near-square overlap stratum where the cue is uninformative reader discrimination was undiminished (0.72 versus the pooled 0.71; Supplementary Table S9), indicating that image content rather than the residual format cue drives the finding; the estimand should nonetheless be read as provenance discrimination together with any residual format attribute. Fifth, readers were a single-country volunteer convenience series, most of them study-group members and group authors of this article, with characteristics beyond expertise tier (age, sex) not collected; because these readers knew the study concerned synthetic-image provenance, their vigilance may have exceeded that of a hypothesis-blind clinical reader, so the reported discrimination is best read as an upper bound on unaided detection; the homogeneous national training background aided expertise-tier classification but limits international generalisability. Sixth, the full-cohort reader variance exceeded the design assumption; equivalence was nonetheless satisfied, and the expertise contrast is reported as bounded within +/−0.10. Seventh, the quality-assurance instrument’s graded axes had limited inter-rater reliability, so the rated-quality association with detection is an exploratory covariate that does not bear on the primary endpoint, for which a three-tier collapsed scale was pre-specified. Finally, a small cluster of pre-data reconciliation steps, conservative with respect to the primary endpoint, and a post-lock correction of the scoring code to its pre-registered monotone form (which moved the pooled area under the curve from 0.66 to 0.71, both within the modest tier, and changed the Faculty–Junior contrast from a directional reading to a non-significant gradient) are disclosed in full in the Supplementary timeline; the encoding error was identified by a structural unit-test collision rather than by any endpoint value, and no arm-level area under the curve or expertise contrast was examined before the final analysis lock.

In a pre-registered, multimodal, confidence-weighted multi-reader multi-case visual Turing test, expert radiologists in a single-country cohort distinguished frontier-generated from authentic radiologic images only modestly: above chance but without a meaningful expertise gradient (equivalence within +/−0.10), with no reader stratum, including board-certified faculty, reaching the pre-specified threshold for a standalone provenance safeguard. These findings motivate radiology AI-literacy efforts and favour provenance-documented learning sources; whether training in synthetic-image cues improves detection is a hypothesis for prospective evaluation, not a result of this study. No provenance-screening claim is licensed by the human-reader area under the curve alone, and unaided expert review is best regarded as one imperfect layer within a broader provenance-integrity framework.

## Data Availability

De-identified, trial-level data (reader responses, confidence ratings, provenance-cue selections, expertise-tier labels, and the locked image-pool manifest, with reader identifiers replaced by non-reversible study codes) will be made available at the Open Science Framework (project DOI 10.17605/OSF.IO/ZRWY8). Individual generated images are available from the corresponding authors for non-commercial research use on reasonable request. No personally identifiable patient data are included.

https://github.com/Yoojin-nam/MeducAI

