## Supplementary Appendix for "Expert Discrimination of AI-Generated versus Authentic Radiologic Images: A Multimodal, Pre-Registered Visual Turing Test"

#### Title

### Supplementary Methods (S1-S21)

*This Supplementary Appendix is self-contained: its reference list (below) is independent of the main manuscript. Section numbers S1-S21 expand the corresponding subsections of the main-text Methods.*

```
correct ~ image_type + expertise_group + image_type:expertise_group
        + modality + subspec + order_index
        + (1 + image_type | reader) + (1 | entity)
```

with image type taking the values authentic, Nano Banana 2, and gpt-image-2, and expertise group taking the values junior, senior, and faculty. The reported quantities are marginal predicted probabilities and odds ratios with 95% confidence intervals; areas under the curve are reported only from the Obuchowski-Rockette/DeLong analyses, not from this model. Because the full random-effects structure requires estimation of an unstructured reader-level covariance matrix under an unbalanced per-reader image-type allocation, singular-fit or non-convergence warnings were anticipated, and a fallback ladder was pre-specified in which the first specification returning a non-singular convergent fit becomes the reported model:

#### Supplementary Table S1. Post-lock timeline: pre-data methodological adjustments, data lock, and the analysis-implementation correction

The image pool and assignment matrix were locked on 2026-05-21. The adjustments below were applied between that lock and the consolidated analysis-plan lock (2026-05-24), all before the 2026-06-05 broadcast and before any reader response was collected. They are disclosed collectively here and referenced in the Discussion Limitations, per the post-lock change-cluster disclosure rule. Per-item artifact identifiers and hashes are archived in the deposited data and code repository.

| # | Date (KST) | Change | Detail and rationale | Timing relative to data |
|---|---|---|---|---|
| 1 | 2026-05-21 | Pre-dispatch lock | Image pool frozen at 241 displayable cells (83 authentic, 78 Nano Banana 2, 80 gpt-image-2; 82 distinct entities; manifest SHA-256 100ef0c0…6558749e). | Pre-broadcast (15 days prior) |
| 2 | 2026-05-21 | Clerical correction | One CT cell's medical-validity flag corrected 0 → 1 (cell retained) on rater self-confirmation; the original record was preserved. | Pre-broadcast; quality-assurance side |
| 3 | 2026-05-21 | Clerical correction | One radiograph cell's medical-validity flag corrected 1 → 0 (cell flagged) on rater self-confirmation. | Pre-broadcast; quality-assurance side |
| 4 | 2026-05-21 | Consensus-fail pool drop | Five Nano Banana 2 cells were dropped under a single prospectively locked rule (corrected medical-validity = 0 and a failed realism gate). | Pre-broadcast; shown to no reader |
| 5 | pre-dispatch | Image-format harmoni | Crop-free, modality-aware normalisation applied symmetrically across all | Pre-broadcast |

| # | Date (KST) | Change | Detail and rationale | Timing relative to data |
|---|---|---|---|---|
| | | sation | three arms. | |
| 6 | pre-dispatch | Authentic-arm view labelling | Nineteen authentic-arm view/specification alignment refinements conservatively labelled as subjective view preference (transparency, not an eligibility gate). | Pre-broadcast |
| 7 | 2026-05-21 | Participant reconciliation | The raw invitee union was reconciled to 61 readers (three acknowledgement-only invitees excluded); historical responses were never edited. | Pre-broadcast |
| 8 | 2026-05-23 | Co-investigator reader exclusion | The pipeline co-investigator was excluded to preserve evaluator–investigator independence (61 → 60 readers; 3,600 assignment rows; matrix SHA-256 2150faa8…87851). | Pre-broadcast; preserves no-data-seen attestation |
| 9 | 2026-05-23 | Measurement-instrument prespecification | The provenance-cue × accuracy contingency analysis was added as a prespecified secondary analysis under a no-data-seen attestation (production responses = 0). | Pre-broadcast; no reader data observed |
| 10 | 2026-05-24 | Analysis prespecifications | An image-quality-axis-dropped quality-restricted variant, a principal-component construct-validity refit, and a false-discovery-rate supplement were added as measurement-instrument prespecifications. | Pre-broadcast |
| 11 | 2026-05-24 | Analysis-plan consolidation | The statistical analysis plan was consolidated into a single canonical document; prior amendment documents were retained as an audit trail. | Pre-broadcast |

| # | Date (KST) | Change | Detail and rationale | Timing relative to data |
|---|---|---|---|---|
| 12 | 2026-06-04 | Pre-registration made public | The preregistration was filed and made public; production responses were verified at zero rows (no-data-seen attestation; OSF DOI 10.17605/OSF.IO/ZRWY8). | One day before broadcast; 0 responses |
| 13 | 2026-06-05 09:00 | Broadcast | The reader task was dispatched; the broadcast date had been advanced across the lock chain so that all quality-assurance–rater readers met the 14-day preferred washout. | t = 0 |
| 14 | data-collection phase | Instrument-failure response recovery | One reader experienced a single technical non-persistence event in which one trial’s response was recorded in the browser but not received by the server, after which the application showed the completion screen and blocked re-access. The reader-confirmed response was incorporated before analysis via a single flagged overlay row; the raw response sheet was not edited. A separate transient recovery was retired as redundant once the original response reached the server. This is collection-phase recovery of a reader’s own response, distinct from the pre-data design changes above. | During collection; reader’s own response |
| 15 | 2026-06-20 | Data lock | The registered window closed with all 60 readers complete (grace window not invoked); the dataset was frozen the next day at 3,600 rows (60 readers × 60 | Post-window-close; no outcome examined before this timestamp |

| # | Date (KST) | Change | Detail and rationale | Timing relative to data |
|---|---|---|---|---|
| | | | trials; dataset SHA-256 d3db6e0e…592ec78). One ineligible investigator-reader's partial records were quarantined and replaced by an eligible faculty reader under the prespecified Faculty-eligibility substitution rule, maintaining 20 Faculty and 60 readers; the recovered overlay was unioned and the pipeline co-investigator (0 rows) excluded. | |
| 16 | 2026-06-20 | Analysis-implementation correction | The pre-registered estimand is the confidence-weighted multi-reader multi-case AUC. The analysis code initially used a non-monotone "folded" encoding of the directional confidence rating that collapsed opposite response categories; it was corrected to the monotone form the registered estimand specifies, identified by a structural unit-test collision rather than by any endpoint value, and the locked analysis was re-run unchanged otherwise (the raw dataset was not modified). The correction moved the primary estimate from 0.66 to 0.71 (both within the modest tier) and changed the expertise contrast from a directional reading to a non-significant gradient with equivalence. | Affects the estimator, not the data |

Between the pre-dispatch lock and the broadcast, a cluster of methodological adjustments was applied: two rater-confirmed clerical corrections, a five-cell consensus-fail drop, crop-free image-format harmonisation, conservative labelling of nineteen authentic-arm view-alignment refinements, participant-list reconciliation, exclusion of the pipeline co-investigator, and three measurement-instrument analysis prespecifications. All preceded any reader response and are itemised with dates above. The five-cell consensus-fail drop removed only low-realism synthetic cells and can therefore only attenuate measured detectability, making it conservative against an above-chance discrimination finding; because these cells were removed before dispatch and shown to no reader, a reader-level re-inclusion analysis is not applicable, and an alternative-rule audit instead documents that the drop predicate (medical-validity = 0 with a failed realism gate) was not tuned to the observed five-cell set, with alternative thresholds removing the same low-realism cells.

#### Supplementary Table S2. MI-CLAIM item-level mapping

Reporting against the MI-CLAIM checklist[23]. Status: **Addressed** / **Partial** / **N/A** (with rationale). This study is a pre-registered reader diagnostic-accuracy evaluation of two commercial text-to-image generators; no predictive model was trained, so MI-CLAIM model-development/optimization/interpretability items are N/A.

| MI-CLAIM part | Item (paraphrased) | Status | Manuscript / Supplement location | Note |
|---|---|---|---|---|
| Part 1. Study design | Clinical problem + intended use stated | Addressed | Introduction; Methods (Study design); Discussion | Provenance discrimination for AI-literacy training; explicitly not a deployable classifier |
| Part 1. Study design | Cohort definition + ground-truth standard | Addressed | Methods (Synthetic images and image pool; Readers) | Reference standard = generation provenance, fixed deterministically at image creation |
| Part 1. Study design | Prospective vs retrospective; registration | Addressed | Methods (Study design, oversight, registration) | Prospective; OSF pre-registered with locked SAP before broadcast |
| Part 2. Data & optimization | Data sources + characteristics | Addressed | Methods (image pool); Suppl Methods S2, S5 | 241 cells / 82 entities / 9 subspecialties / 6 modalities; authentic = teaching repository |

| MI-CLAIM part | Item (paraphrased) | Status | Manuscript / Supplement location | Note |
|---|---|---|---|---|
| Part 2. Data & optimization | Preprocessing / transformations | Addressed | Methods (normalization); Suppl Methods S4 | Single crop-free, modality-aware normalization applied symmetrically across arms |
| Part 2. Data & optimization | Train/ validation/test partition + independence | N/A (model dev) | — | No model trained; prompt development used a frame-external development set (Suppl Methods S3), kept independent of the locked study frame |
| Part 2. Data & optimization | Class balance / cohort selection | Addressed | Methods (image pool; assignment matrix) | Balanced, arm-balanced incomplete reader-image assignment: per reader 20 authentic / 20 Nano Banana 2 / 20 gpt-image-2, no entity repeated |
| Part 3. Model performance | Primary performance metric + uncertainty | Addressed | Methods (Statistical analysis); Results | Confidence-weighted MRMC AUC with 95% CI (Obuchowski-Rockette/DeLong) |
| Part 3. Model performance | Comparison to a meaningful baseline | Addressed | Methods; Results | Tested against chance (0.5); expertise-stratified contrast under TOST equivalence |

| MI-CLAIM part | Item (paraphrased) | Status | Manuscript / Supplement location | Note |
|---|---|---|---|---|
| Part 4. Model examination | Interpretability / explanation technique | Partial | Methods (provenance-cue analysis); Results | No model internals; instead a prespecified human-reported provenance-cue × accuracy analysis |
| Part 4. Model examination | Sensitivity / subgroup analyses | Addressed | Methods (Sensitivity analyses); Suppl Methods S9-S16 | Four pre-specified sensitivity analyses incl. quality-restricted and aspect-ratio-stratified |
| Part 5. Reproducibility | Code availability | Addressed | Methods (Data and code availability); Suppl Methods S18-S19 | Analysis code archived on Zenodo (DOI at publication); pipeline at github.com/Yoojin-nam/MeducAI (Apache-2.0) |
| Part 5. Reproducibility | Data availability | Addressed | Methods (Data and code availability) | De-identified reader data + normalized-image hashes + assignment matrix on Zenodo (CC BY 4.0) |
| Part 5. Reproducibility | Model / environment availability | Partial / N/A | Suppl Methods S2-S3 | Generators are commercial APIs frozen at a model snapshot; frozen prompts + model identifiers + per-image SHA-256 provided; vendor model weights not author- |

| MI-CLAIM part | Item (paraphrased) | Status | Manuscript / Supplement location | Note |
|---|---|---|---|---|
| | | | | controlled |

#### Supplementary Table S3. Diversity and representativeness

Reader-cohort and image-pool diversity are summarized below, per NEJM AI reporting requirements for clinical research.

##### Reader cohort (n = 60 dispatched)

| Dimension | Categories | n (as dispatched) | Note |
|---|---|---|---|
| Expertise tier | Faculty (board-certified) / Senior (PGY 3-4) / Junior (PGY 1-2) | 20 / 18 / 22 | All 60 completed (Faculty 20 / Senior 18 / Junior 22); binding contrast Faculty (20) vs Junior (22) |
| Institution | Multi-center (>30 institutions across named authors and the reader study group) | multi-center | Predominantly tertiary academic centers; affiliations listed in the byline and the study-group appendix |
| Subspecialty | Multiple radiologic subspecialties (faculty subspecialty recorded at enrollment) | faculty-recorded | Reader subspecialty was collected for faculty; not used as a primary contrast |
| Geography | Republic of Korea (60) | 60 | Single-country reader cohort; international generalizability is a stated limitation |
| Sex / age | Not systematically collected | not collected | Reader demographics beyond expertise tier were not collected; reported as a limitation |

##### Image pool (241 displayable cells; 82 entities)

| Dimension | Categories | n | Note |
|---|---|---|---|
| Arm | Authentic / Nano Banana 2 / gpt-image-2 | 83 / 78 / 80 | Locked image-pool manifest |

| Dimension | Categories | n | Note |
|---|---|---|---|
| Modality | 6 modalities | CT 99, MR 67, US 33, radiograph 25, mammography 9, angiography 8 | Multimodal scope distinguishes from single-modality prior work |
| Subspecialty | 9 subspecialties | abdominal 27, breast 30, cardiovascular 25, genitourinary 27, interventional 26, musculoskeletal 26, neuro/head-and-neck 27, pediatric 27, thoracic 26 | Cross-subspecialty coverage |
| Patient demographics (authentic arm) | Sex / age / race | Not available | Teaching-repository source lacks consistent per-image demographics; patient-demographic fairness cannot be assessed (stated limitation; paired with the fairness-reporting item) |

Per-modality and per-subspecialty image-pool counts computed from the locked image-pool manifest; reader tier composition from the final analysis cohort (all 60 completed). These are design/coverage descriptors, not study outcomes.

### Supplementary Results

All quantities are recomputed from the locked dataset (SHA-256 d3db6e0e…592ec78) by the committed analysis scripts; see the provenance manifest. The primary confidence-weighted, reader-averaged area under the curve (AUC) conditional on the locked image pool was 0.71 (95% CI, 0.69 to 0.74).

#### Supplementary Table S4. Pre-specified sensitivity analyses (primary endpoint)

| Analysis | AUC or estimate | 95% CI | Δ vs primary | Divergence trigger | Conclusion |
|---|---|---|---|---|---|
| Quality-restricted AUC (QA-pass synthetic + matched | 0.686 | (0.658 to 0.716) | -0.028 | ≥0.05: False | concordant; not deployable |

| Analysis | AUC or estimate | 95% CI | Δ vs primary | Divergence trigger | Conclusion |
|---|---|---|---|---|---|
| real) | | | | | |
| Quality-restricted AUC, image-quality-axis-dropped variant | 0.689 | (0.658 to 0.717) | 0.003 vs quality-restricted | ≥0.03: False | robust to lowest-reliability axis |
| Complete-coverage entities (75/82) | 0.719 | (0.692 to 0.746) | 0.005 | ≥0.02: False | per-model coverage not a confounder |
| Quality-assurance–rater exclusion (n=54, Faculty 14) | 0.709 | (0.682 to 0.736) | -0.005 | interpretive | gradient direction robust |
| Reader-and-case generalization bootstrap (supplementary) | 0.714 | (0.675 to 0.751) | — | — | above chance with case variance |

PI-realism stratification was vacuous (all 241 locked cells PI-pass). Washout was vacuous (11/11 quality-assurance faculty ≥14 days at broadcast). Abstention was vacuous (all 60 readers 60/60). Fast-click audit: 0.25% of responses <2 s (n=9), descriptive only. Reader-cluster bootstrap = 2,000 iterations, seed 42.

#### Supplementary Table S5. Quality-assurance image quality and reader detection (synthetic-only GLMM)

| Predictor | Odds ratio | 95% CI | Interpretation |
|---|---|---|---|
| QA 5-axis composite (z-sum) | 0.843 | (0.797 to 0.893) | higher QA quality → lower odds of detection |
| QA composite (PCA factor-1, construct-validity refit) | 0.837 | — | Cronbach α 0.823; Δ 0.7% vs z-sum (no divergence) |

Synthetic cells only; outcome = correctly identifying an AI image as AI. Random effects: (1 + image_type | reader) + (1 | entity).

#### Supplementary Table S6. Quality-assurance instrument reliability

Calibration-round 5-point Krippendorff α ranged 0.04–0.39 across the five graded axes (pre-data); the 3-tier collapsed scale is the primary reporting scale for axes below 0.50. Second-reader agreement (ICC(3,1), two-way absolute, single measures; n=60 paired cells):

| Graded axis | ICC(3,1) |
|---|---|
| modality_accuracy | 0.109 |
| main_finding | 0.242 |
| anatomical_accuracy | 0.329 |
| image_quality | 0.100 |

Medical-validity agreement was complete (both raters rated all paired cells valid; κ undefined for lack of variance). Graded-axis ICCs were low, consistent with the calibration-round α; the QA composite enters the analysis only as a covariate (Supplementary Table S5), and the 3-tier collapsed scale is used for primary reliability reporting.

#### Supplementary Table S7. Per-modality and per-subspecialty detection (descriptive)

| Stratum | AUC | 95% CI |
|---|---|---|
| Angio | 0.682 | (0.523 to 0.818) |
| CT | 0.714 | (0.673 to 0.754) |
| Mammo | 0.703 | (0.562 to 0.828) |
| MRI | 0.681 | (0.639 to 0.723) |
| US | 0.789 | (0.742 to 0.834) |
| XR | 0.626 | (0.568 to 0.685) |
| abdominal (subspecialty) | 0.702 | (0.629 to 0.773) |
| breast (subspecialty) | 0.747 | (0.704 to 0.790) |
| cardiovascular (subspecialty) | 0.680 | (0.619 to 0.741) |
| gu (subspecialty) | 0.689 | (0.623 to 0.755) |
| interventional (subspecialty) | 0.752 | (0.689 to 0.813) |
| msk (subspecialty) | 0.640 | (0.572 to 0.709) |
| neuro_hn (subspecialty) | 0.757 | (0.703 to 0.809) |
| pediatric (subspecialty) | 0.741 | (0.686 to 0.793) |
| thoracic (subspecialty) | 0.719 | (0.658 to 0.775) |

Reader-cluster bootstrap 95% CI; descriptive (not powered for stratified inference).

#### Supplementary Table S8. Post-hoc exploratory analyses (descriptive)

**Confidence calibration.** Accuracy by reported confidence level:

| Confidence | n | Accuracy | 95% CI (Wilson) |
|---|---|---|---|
| 1 | 107 | 0.551 | (0.457 to 0.642) |
| 2 | 401 | 0.584 | (0.535 to 0.631) |
| 3 | 1008 | 0.626 | (0.596 to 0.655) |
| 4 | 1601 | 0.693 | (0.670 to 0.715) |
| 5 | 483 | 0.832 | (0.796 to 0.863) |

Mean confidence on errors 3.34 vs correct 3.64; 27.5% of high-confidence (4–5) judgements were incorrect.

**Between-reader variation.** Per-reader AUC mean 0.714, SD 0.107, range 0.449 to 0.939; 3 of 60 readers at or below chance.

**Response time** (exploratory, descriptive). Median 12.9 s (IQR 8.2–22.3 s; 99th percentile 104.3 s). Multi-session re-access (consecutive same-assignment trial gap ≥1 h) and extreme reconnect anomalies are described in the analysis notes; no response was excluded from the endpoint.

**Lowest-accuracy entities.** The synthetic entities least often identified clustered in ultrasound, radiography, and mammography (lowest entity accuracy 0.226).

#### Supplementary Table S9. Aspect-ratio confound bound and de-confounded reader detection

Native aspect ratio was the one residual format attribute not matched across arms. A univariate deviation-from-square classifier bounds how much the format cue alone can separate the arms (the separability ceiling); reader detection is then re-estimated within the near-square overlap stratum where the cue is uninformative.

| Quantity | AUC | Note |
|---|---|---|
| Univariate aspect-ratio classifier, full pool (separability ceiling) | 0.701 | format-only upper bound |
| Univariate aspect-ratio classifier, near-square overlap stratum | 0.576 | ~0.5 = format uninformative here |
| Reader-averaged confidence-weighted AUC, full pool | 0.714 | exceeds the format ceiling |
| Reader-averaged confidence-weighted AUC, near-square overlap stratum (de-confounded) | 0.716 | format cue uninformative |

| Quantity | AUC | Note |
|---|---|---|
| Attenuation (full pool − overlap stratum) | -0.002 | negligible |

Near-square overlap stratum: aspect ratio in [1.000, 1.340] (154 AI cells, 54 authentic cells); reader-averaged over 60 readers. Reader detection exceeds the format-only ceiling and is undiminished where the cue is uninformative, indicating that image content rather than the residual format attribute drives detection; the estimand should nonetheless be read as provenance discrimination together with any residual format attribute.

#### Appendix: Members of the MeducAI Reader Study Group

The 44 members of the MeducAI Reader Study Group, who served as readers in the visual Turing test and are group authors of this article, are listed below. Each member completed the reader task under electronic informed consent and consented to group authorship at enrollment, and is listed as a collaborator for bibliographic indexing.

Jeonghyeon Ahn, MD[1]; Junho An, MD[2]; Hye Ree Cho, MD[3]; Soong Moon Cho, MD[4]; Jun Young Choi, MD[5]; Won Jae Choi, MD[6]; Kyung Rae Do, MD[7]; Inae Hwang, MD[3]; Jun Yong Im, MD[8]; Hwi Young Jang, MD[8]; Dongjun Jeong, MD[9]; Sangwoo Joh, MD[10]; Yohan Joo, MD[11]; Sunghyun Jung, MD[12]; Hyeon Wook Kang, MD[13]; Chaerin Kim, MD[14]; Donghyun Kim, MD[12]; Eunji Kim, MD[3]; Hyunsoo Kim, MD[15]; Jaeyoon Kim, MD[3]; Kyubin Kim, MD[7]; Seung Kwan Kim, MD[16]; Yeonsu Kim, MD[3]; Ohmin Kwon, MD[17]; Hwa Jin Lee, MD[18]; Hye-Won Lee, MD[19]; Hyeyun Lee, MD[7]; Joonhyuk Lee, MD[13]; Kanghwi Lee, MD[12]; Kyung Eun Lee, MD[20]; Noha Lee, MD[21]; Suji Lee, MD[22]; Yoonhee Lee, MD[10]; Sung Gong Lim, MD[5]; Suam Oh, MD[22]; Dawon Park, MD[23]; Ho Young Park, MD[24]; Jae Eun Park, MD[1]; Jiho Park, MD[15]; Kyeong Jin Park, MD[13]; Seo Ha Park, MD[25]; Yoojin Park, MD[10]; Kyeong Hwa Ryu, MD[3]; Dongmin Shin, MD[1].

##### Affiliations

1. Department of Radiology, Nowon Eulji Medical Center, Seoul, Republic of Korea
2. Department of Radiology, SMG Yonsei Hospital, Changwon, Republic of Korea
3. Department of Radiology, Samsung Changwon Hospital, Sungkyunkwan University School of Medicine, Changwon, Republic of Korea
4. Department of Radiology, Heung-K Hospital, Siheung, Republic of Korea
5. Department of Radiology, Gyeongsang National University Changwon Hospital, Changwon, Republic of Korea
6. Department of Radiology, Bobath Memorial Hospital, Hanam, Republic of Korea
7. Department of Radiology and Research Institute for Convergence of Biomedical Science and Technology, Pusan National University Yangsan Hospital, Pusan National University School of Medicine, Yangsan, Republic of Korea
8. Department of Radiology, Naeun Hospital, Incheon, Republic of Korea
9. Department of Radiology, Daegu Catholic University Medical Center, Daegu, Republic of Korea
10. Department of Radiology, Gil Medical Center, Gachon University College of Medicine, Incheon, Republic of Korea

11. Department of Radiology and Research Institute of Radiological Science, Severance Hospital, Yonsei University College of Medicine, Seoul, Republic of Korea
12. Department of Radiology, Seoul National University Hospital, Seoul National University College of Medicine, Seoul, Republic of Korea
13. Department of Radiology, Seoul National University Bundang Hospital, Seongnam, Republic of Korea
14. Department of Radiology, Ilsan Paik Hospital, Inje University College of Medicine, Goyang, Republic of Korea
15. Department of Radiology, Seoul St. Mary's Hospital, College of Medicine, The Catholic University of Korea, Seoul, Republic of Korea
16. Department of Radiology, Kangnam General Hospital, Yongin, Republic of Korea
17. Department of Radiology, Yeouido St. Mary's Hospital, The Catholic University of Korea, Seoul, Republic of Korea
18. Department of Radiology and Research Institute of Radiology, University of Ulsan College of Medicine, Asan Medical Center, Seoul, Republic of Korea
19. Department of Radiology, Yonsei Best Hospital, Uijeongbu, Republic of Korea
20. Department of Radiology, Korea University Guro Hospital, Korea University College of Medicine, Seoul, Republic of Korea
21. Department of Radiology, Chosun University Hospital and Chosun University College of Medicine, Gwangju, Republic of Korea
22. Department of Radiology, Samsung Medical Center, Sungkyunkwan University School of Medicine, Seoul, Republic of Korea
23. Department of Radiology, CHA Bundang Medical Center, Seongnam, Republic of Korea
24. Department of Radiology, Yeson Hospital, Bucheon, Republic of Korea
25. Department of Radiology, Gangneung Asan Hospital, University of Ulsan College of Medicine, Gangneung, Republic of Korea
1 Nam Y *et al.* Human-in-the-Loop Validation of a Sequential Multi-LLM Medical Education Pipeline. 2026.

2 Nam Y *et al.* Paper 2 OSF Preregistration v23 — Visual Turing Test of Synthetic Radiologic Image Provenance. 2026. DOI:10.17605/OSF.IO/ZRWY8.

3 Tordjman M, Yuce M, Ammar A, *et al.* The Rise of Deepfake Medical Imaging: Radiologists' Diagnostic Accuracy in Detecting ChatGPT-generated Radiographs. *Radiology* 2026; **318**: e252094.

4 Krippendorff K. Content Analysis: An Introduction to Its Methodology, 4th edn. Thousand Oaks, CA: SAGE Publications, 2018 DOI:10.4135/9781071878781.

5 Hallgren KA. Computing Inter-Rater Reliability for Observational Data: An Overview and Tutorial. *Tutorials in quantitative methods for psychology* 2012; **8**: 23–34.

6 Hillis SL. A comparison of denominator degrees of freedom methods for multiple observer ROC analysis. *Statistics in medicine* 2007; **26**: 596–619.

7 Obuchowski NA. Multireader receiver operating characteristic studies: a comparison of study designs. *Acad Radiol* 1995; **2**: 709–16.

8 Beam CA, Layde PM, Sullivan DC. Variability in the interpretation of screening mammograms by US radiologists. Findings from a national sample. *Arch Intern Med* 1996; **156**: 209–13.

9 Schuirmann DJ. A comparison of the two one-sided tests procedure and the power approach for assessing the equivalence of average bioavailability. *J Pharmacokinet Biopharm* 1987; **15**: 657–80.

10 Lakens D. Equivalence tests: A practical primer for t tests, correlations, and meta-analyses. *Soc Psychol Personal Sci* 2017; **8**: 355–62.

11 Smith BJ, Hillis SL, Pesce LL. MRMCaov: Multi-Reader Multi-Case Analysis of Variance. 2025. DOI:10.32614/CRAN.package.MRMCaov.

12 DeLong ER, DeLong DM, Clarke-Pearson DL. Comparing the areas under two or more correlated receiver operating characteristic curves: a nonparametric approach. *Biometrics* 1988; **44**: 837–45.

13 Bates D, Mächler M, Bolker B, Walker S. Fitting Linear Mixed-Effects Models Using lme4. *J Stat Softw* 2015; **67**. DOI:10.18637/jss.v067.i01.

14 Brooks ME, Kristensen K, Benthem KJ van, *et al.* glmmTMB Balances Speed and Flexibility Among Packages for Zero-inflated Generalized Linear Mixed Modeling. *R J* 2017; **9**: 378.

15 Sounderajah V, Ashrafian H, Aggarwal R, *et al.* Developing specific reporting guidelines for diagnostic accuracy studies assessing AI interventions: The STARD-AI Steering Group. *Nat Med* 2020; **26**: 807–8.

16 Bossuyt PM, Reitsma JB, Bruns DE, *et al.* STARD 2015: an updated list of essential items for reporting diagnostic accuracy studies. *BMJ* 2015; **351**: h5527.

17 Tejani AS, Klontzas ME, Gatti AA, *et al.* Checklist for Artificial Intelligence in Medical Imaging (CLAIM): 2024 Update. *Radiol Artif Intell* 2024; **6**: e240300.

18 Mongan J, Moy L, Kahn CE. Checklist for Artificial Intelligence in Medical Imaging (CLAIM): A Guide for Authors and Reviewers. *Radiol Artif Intell* 2020; **2**: e200029.

19 Park SH, Suh CH, Lee JH, *et al.* Minimum Reporting Items for Clear Evaluation of Accuracy Reports of Large Language Models in Healthcare (MI-CLEAR-LLM): 2025 Updates. *Korean journal of radiology* 2025; **26**: 1123–32.

20 Gallifant J, Afshar M, Ameen S, *et al.* The TRIPOD-LLM reporting guideline for studies using large language models. *Nat Med* 2025; **31**: 60–9.

21 Servier Laboratories. SMART Servier Medical Art. 2024. https://smart.servier.com/.

22 Nam Y, An T, Hwang SI, *et al.* Feasibility and psychometric evaluation of large language model–generated radiology learning materials for board examination preparation: a prospective pilot study. *BMC Med Educ* 2026. DOI:10.1186/s12909-026-09704-8.

23 Norgeot B, Quer G, Beaulieu-Jones BK, *et al.* Minimum information about clinical artificial intelligence modeling: The MI-CLAIM checklist. *Nat Med* 2020; **26**: 1320–4.
